# Reconstructing real-world metastatic lines of therapy enables progression risk stratification in breast cancer

**DOI:** 10.64898/2026.02.24.26346242

**Authors:** Xuan Zhao, Thibault Niederhauser, Zsolt Balázs, Andreas Wicki, Bowen Fan, Michael Krauthammer

## Abstract

Estimating progression-free survival (PFS) for each metastatic line of therapy (mLoT) is valuable yet challenging in precision oncology, as both are rarely available as paired labels. We present (i) an evidence enrichment pipeline that reconstructs mLoTs and PFS from longitudinal EHR and (ii) a model for progression risk prognostication. In the MSK-CHORD breast cancer cohort, the pipeline yielded 8,791 mLoTs from 2,881 patients, and the trained model achieved Antolini’s C **= 0.681 *±* 0.006** and integrated Brier score (IBS) **= 0.124 *±* 0.004** in nested cross-validation. We validated our pipeline by reconstructing regimen and PFS labels in the AACR GENIE BPC cohorts from DFCI and VICC. Of the reconstructed regimens, 86.2% and 82.4% matched a curated regimen, with PFS event agreement 93.2% and 94.6% among matches. On the curated BPC regimens, the MSK-CHORD model achieved C **= 0.649*±*0.002** and **0.634*±*0.006**, with IBS **= 0.166 *±* 0.001** and **0.159 *±* 0.001**, despite several features of original model missing. The model remains robust to modality ablation and stratifies prognosis interpretably despite substantial cohort imbalance and heterogeneity. Together, this supports scalable characterization of metastatic-line PFS across heterogeneous regimens and hypothesis generation, pending prospective clinical validation.

## Introduction

Metastatic breast cancer (mBC) is an advanced disease typically treated with successive systemic lines of therapy (metastatic lines of therapy; mLoTs) [1, 2], with the goals of controlling disease, prolonging survival, and maintaining quality of life. Line-of-therapy efficacy is often quantified using progression-free survival (PFS), defined as time from treatment initiation to disease progression or death [3]. Prior work on mBC PFS prediction has often focused on curative or first-line metastatic therapy [4], or on specific regimens or cohorts. For example, Varma et al. [5] used natural language processing (NLP) to derive PFS for patients treated with endocrine therapy plus a CDK4/6 inhibitor; Merzhevich et al. [6] modeled PFS in a ribociclib-treated cohort; and Cabel et al. [7] studied later-line outcomes in triple-negative breast cancer (TNBC). Beyond first line (mLoT2+), treatment benefits often diminish, while regimens become increasingly heterogeneous [8–10], making PFS evidence acquisition more difficult.

An interesting exploration is therefore to model PFS across heterogeneous metastatic treatment lines using diverse real-world data. Although controlled, homogeneous PFS and mLoT pairs are sparse, longitudinal electronic health records (EHR) can be mined for consecutive, heterogeneous mLoTs with credible progression endpoints. This creates an opportunity to share information across population subtypes and heterogeneous treatment regimens while restricting analysis to episodes that meet specified reconstruction and PFS-labeling rules. Developing such a framework has two practical challenges. First, clinically coherent mLoT reconstruction and credible PFS labeling are difficult in routine data: line boundaries and segmentation algorithms vary across cancer types, data sources, and conventions [11–14]. Progression is rarely recorded as a structured endpoint and is often inferred from unstructured documentation, increasingly via NLP approaches [5, 15–17]. Second, rich multimodal predictors can induce leakage and shortcut learning; for example, complete within-line treatment records can encode post-initiation adaptations, such as switches for toxicity or early progression, and extensive surveillance patterns can proxy for poor prognosis.

In this work, we address both challenges and propose a novel framework to model mBC PFS across heterogeneous, successive metastatic lines of therapy. Our two-step workflow (Fig. 1a) proceeds as follows: First, we reconstruct mLoTs with clinically informed heuristics, apply stringent filtering, and anchor PFS to previously validated MSK-CHORD NLP-derived radiology progression assessments, prioritizing label fidelity and clinical plausibility over maximum cohort size. Second, we use these reconstructed PFS endpoints to model line-specific progression risk with machine learning models, enforcing rules to reduce leakage from subsequent treatment adaptations and non-biological proxies.

**Fig. 1:**
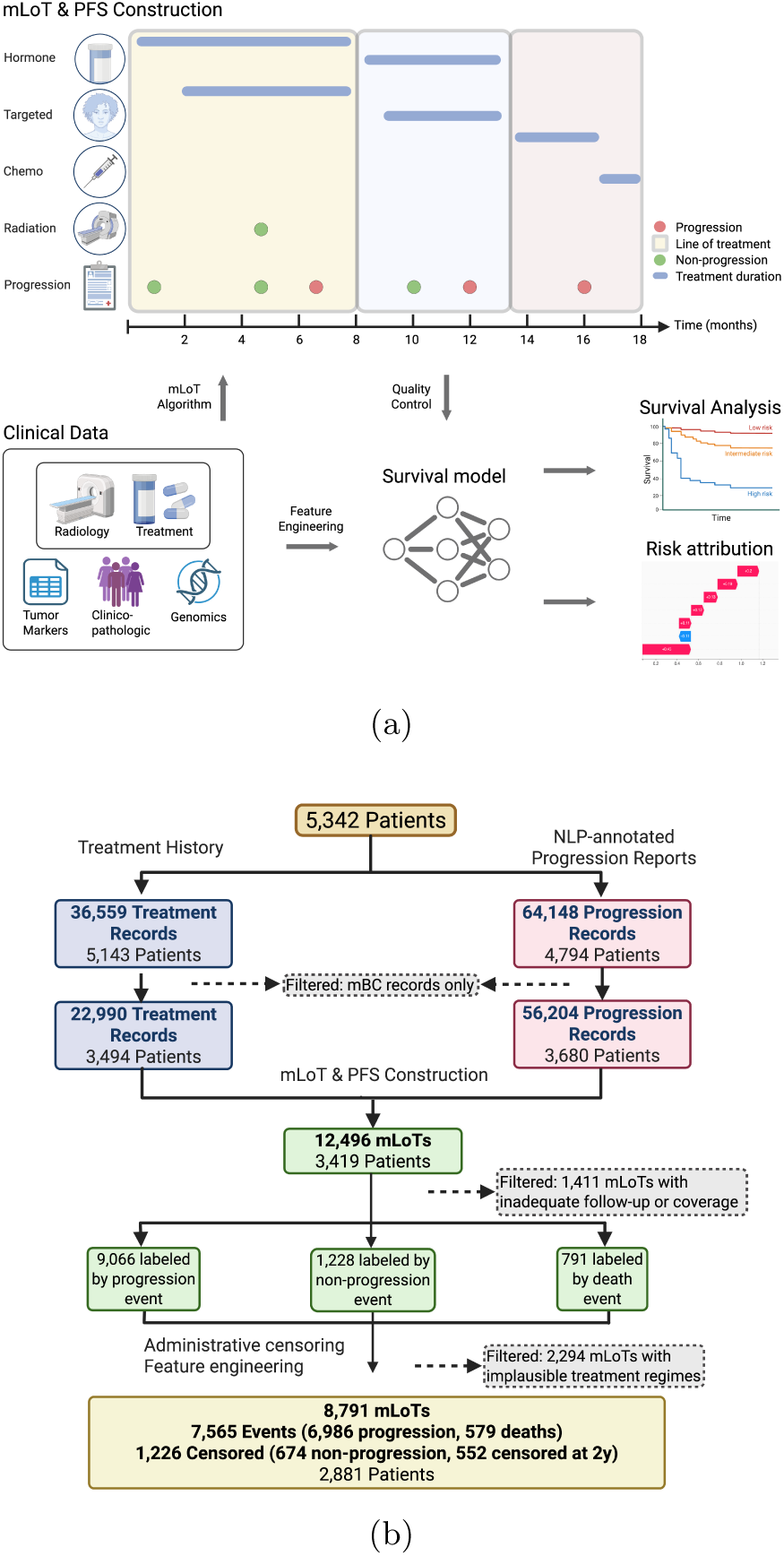
Workflow overview. **a**: The workflow combines NLP-annotated radiology reports and treatment history to segment mLoTs and assign PFS outcomes. Survival models are trained on five clinical modalities, with outputs evaluated for discrimination, calibration, interpretability, and robustness. **b**: Derivation of the analytic mLoT cohort and inclusion criteria. Both created with BioRender [43]. Created in BioRender. Zhao, X. (2026) https://BioRender.com/56dcnu2.

We retrospectively reconstructed metastatic treatment episodes to yield line-level PFS evidence from tabular data in MSK-CHORD. We validated our mLoT algorithm and trained model separately on GENIE BPC’s Dana-Farber Cancer Institute (DFCI) and Vanderbilt-Ingram Cancer Center (VICC) cohorts [18, 19]. 82–86% of derived regimens match existing curated regimens, and matches plus grouping differences from deliberate design choices account for 94.3% and 94.2%. Among matched regimen pairs, PFS event indicators agreed in 93.2% at DFCI and 94.6% at VICC. Risk ranking transferred partly to both external cohorts. A patient-level cross-validated local update reduced external calibration error while the underlying GBSA models remained fixed. Our model supports interpretable progression-risk stratification and an extent of transferability.

## Results

### Cohort overview

We applied our evidence enrichment pipeline to the mBC subset of the MSK-CHORD dataset [17]. The resulting analytic cohort includes 2,881 patients contributing 8,791 mLoTs (Table 1; Supplementary Table 1). HR+/HER2*−* disease comprises 67.6% of patients, followed by 15.3% triple-negative breast cancer (TNBC) and 17.1% HER2+. mLoT1 shows lower event rates than the population average, a result of both the predominance of mLoT2+ episodes (73.4%) and diminishing benefits of therapy. TNBC mLoTs have higher event rates, shorter median PFS, and fewer lines received than other subtypes, plausibly reflecting fewer effective treatment options and a more severe prognosis. First-line treatment patterns differed by subtype, with chemotherapy dominant in TNBC, HER2-directed therapy prominent in HER2-positive disease, and endocrine-based therapy dominant in HR+/HER2*−* disease (Supplementary Table 2). In later lines, subtype-specific differentiation is partially attenuated and broadly applicable regimen categories recur across groups (Supplementary Table 3).

**Table 1:** Cohort size, PFS summary, and first-line (mLoT1) treatment-category combinations overall and by disease subtype. Median PFS is Kaplan–Meier estimated (days); event rate is percent.

| **a. Cohort characteristics** |  |  |  |  |  |
| --- | --- | --- | --- | --- | --- |
|  | Overall | TNBC | HR–<br>HER2+ | HR+<br>HER2– | HR+<br>HER2+ |
| Patients | 2,881 | 440 | 147 | 1,948 | 346 |
| Total mLoTs | 8,791 | 1,032 | 437 | 6,130 | 1,192 |
| Mean mLoTs/patient | 3.05 | 2.35 | 2.97 | 3.15 | 3.45 |
| Event rate (%) | 86.1 | 91.1 | 76.0 | 86.5 | 83.3 |
| Median PFS (days) | 115 | 85 | 155 | 116 | 145 |
| First lines | 2,342 | 389 | 141 | 1,565 | 247 |
| First-line event rate (%) | 71.1 | 83.3 | 63.1 | 69.3 | 67.6 |
| Top 1 mLoT1 |  | Chemo | Chemo,<br>Biologic | Hormone,<br>Targeted | Chemo,<br>Biologic |
| – # (%) |  | 289 (74.3%) | 81 (57.4%) | 703 (44.9%) | 82 (33.2%) |
| Top 2 mLoT1 |  | Chemo,<br>Immuno | Biologic | Hormone | Hormone,<br>Biologic |
| – # (%) |  | 39 (10.0%) | 25 (17.7%) | 528 (33.7%) | 55 (22.3%) |

### Agreement of mLoT reconstruction with curated GENIE BPC regimens

To assess reconstruction agreement, we applied our algorithm to the GENIE BPC dataset, and compared against existing curated regimens and outcome labels. We use BPC’s PFS-I and Mixed labels [20] to indicate presence of progression, which matches the positive class of the MSK-CHORD NLP-derived progression labels. Using BPC data, 646/749 (86.2%) reconstructed episodes at DFCI and 314/381 (82.4%) at VICC matched an existing BPC regimen. A design choice to note is that BPC records a treatment break of at least eight weeks as the end of a regimen [20], whereas our algorithm advances a line after qualifying progression or a qualifying new drug started at least one year after line start (Methods, “mLoT segmentation”). In total, 706/749 (94.3%) reconstructed episodes at DFCI and 359/381 (94.2%) at VICC either matched a BPC regimen or combined BPC regimens after treatment changes without counted progression (Supplementary Table 19). Among the 646 DFCI and 314 VICC matched pairs, event indicators agreed in 93.2% and 94.6%, while both event status and exact PFS duration agreed in 519/646 (80.3%) and 265/314 (84.4%), respectively (Supplementary Table 19b). Including differences in design choices (Methods, “mLoT segmentation” and “PFS labeling and censoring”), 623/646 (96.4%) and 307/314 (97.8%) of matched pairs either agreed exactly or had a listed endpoint-rule difference (Supplementary Table 19).

### Model performance and calibration

We compared five model families spanning proportional hazards (PH [21]) and non-PH assumptions and linear, tree-based, and deep learning approaches under nested cross-validation (nested CV). Model outputs are line-specific PFS probability curves over a 2-year horizon, and model class does not materially affect performance in this setting. A gradient-boosted survival model with CoxPH loss and regression trees as base learners from scikit-survival (GBSA [22]) achieves the near-best global C-index and mean AUC, as well as the best global integrated Brier score (IBS) and horizon-specific AUC (Table 2). Pooled out-of-fold GBSA C was 0.6807 (95% CI 0.6741–0.6873) and remained 0.6807 after comparisons between lines from the same patient were excluded (Supplementary Tables 13 and 22).

**Table 2:** Global performance across survival models for PFS prediction. C-index: Antolini’s C [39]. Mean AUC: weighted mean of the integrated cumulative/dynamic time-dependent area under the ROC curve (cdAUC [40]). AUC @ 1y and AUC @ 2y: cdAUC at 1 and 2 years. IBS: integrated Brier score [42]. Values are mean *±* SD across five outer test folds, using all 8,791 retained mLoTs from 2,881 patients. Pooled out-of-fold estimates and first-line estimates are reported separately.

| Metric | CoxPH | DeepHit | DeepSurv | GBSA | RSF |
| --- | --- | --- | --- | --- | --- |
| C-index | 0.679 ± 0.007 | **0.685 ± 0.008** | 0.680 ± 0.009 | 0.681 ± 0.006 | 0.674 ± 0.006 |
| mean AUC | 0.754 ± 0.008 | **0.759 ± 0.007** | 0.755 ± 0.011 | 0.757 ± 0.006 | 0.747 ± 0.005 |
| AUC @ 1y | 0.818 ± 0.008 | 0.811 ± 0.008 | 0.816 ± 0.012 | **0.824 ± 0.008** | 0.814 ± 0.009 |
| AUC @ 2y | 0.842 ± 0.013 | 0.806 ± 0.026 | 0.841 ± 0.015 | **0.847 ± 0.012** | 0.845 ± 0.012 |
| IBS | 0.126 ± 0.004 | 0.140 ± 0.007 | 0.125 ± 0.005 | **0.124 ± 0.004** | 0.125 ± 0.004 |

Time-dependent performance varied more by evaluation horizon than by model class, with increasing cdAUC and decreasing IPCW C over follow-up (Fig. 2a; Supplementary Table 4). Full per-horizon IPCW-*C*, AUC, and Brier scores are reported in Supplementary Table 4; subtype-stratified curves exhibit wider uncertainty in less prevalent groups (Supplementary Figure 1). Across both metrics, DeepHit shows an additional decline in the final 180 days of follow-up, suggesting less stable long-term performance and not ideal for downstream analysis. Because downstream analyses prefer stable risk ranking and calibrated survival curves, we prioritized IBS, horizon stability, and interpretability over the marginally higher global C-index of DeepHit and selected GBSA for post-hoc analysis.

**Fig. 2:**
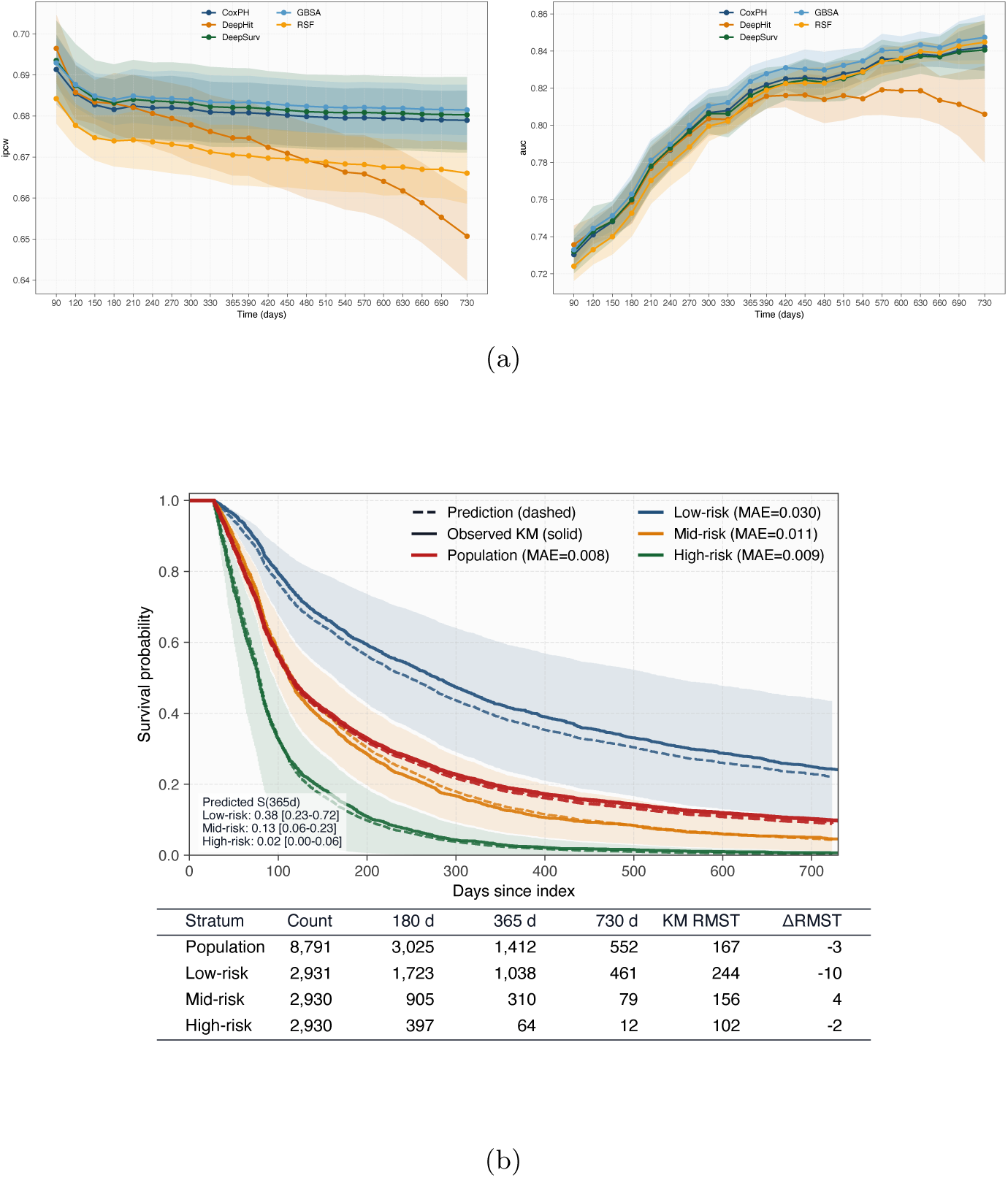
Time-dependent model performance and population risk stratification. **a**: IPCW C-index (left) and cdAUC (right) across evaluation horizons (90 d–2 y) for all models. Curves show means with shaded *±*1 SD across nested CV outer folds. **b**: Disease subtype population (red) and tertile-stratified (blue, orange, and green) Kaplan–Meier (solid) and mean predicted survival (dashed). Shaded area indicates 95% predicted curves within each tertile. Table shows at-risk counts and RMST summaries. RMST@365: KM RMST with cutoff at 365 days. Δ RMST: predicted RMST - KM RMST. Left legend: range of survival probability at 365 days. Reported 2-year (730 d) summaries are evaluated at the closest available time point on the evaluation grid and labeled as 730 d.

We measured calibration with the integrated calibration index (ICI [23]), which was 1.75, 1.57, and 0.91 percentage points at 180, 365, and 730 days (Supplementary Table 25a), respectively. This means that on average, a regimen’s predicted probability of progression by each horizon differed by less than 2 percentage points from the observed probability among regimens with similar predictions.

We compared the multimodal models with a baseline Cox reference using first-line status, HR and HER2 status, the most recent ECOG value, liver involvement in the preceding 90 days, and log transformed most recent CA15–3 value. In MSK-CHORD, C increased from 0.6426 to 0.6782 for full CoxPH and 0.6807 for full GBSA, with paired increases of 0.0355 (95% CI 0.0305–0.0405) and 0.0380 (0.0331–0.0429), respectively; IBS decreased from 0.1343 to 0.1257 and 0.1241 (Supplementary Table 23). The full feature set increased discrimination above chance, by approximately 25% and 27%, respectively. Removing any single clinical modality (Supplementary Note 1 Feature variables) results in only modest performance degradation (Fig. 3b). Among single-modality models (Fig. 3a), imaging-derived features capturing disease burden and metastatic sites provide the strongest standalone discrimination, with planned regimen features performing similarly. By contrast, genomics and clinicopathologic features provide weaker standalone signal.

**Fig. 3:**
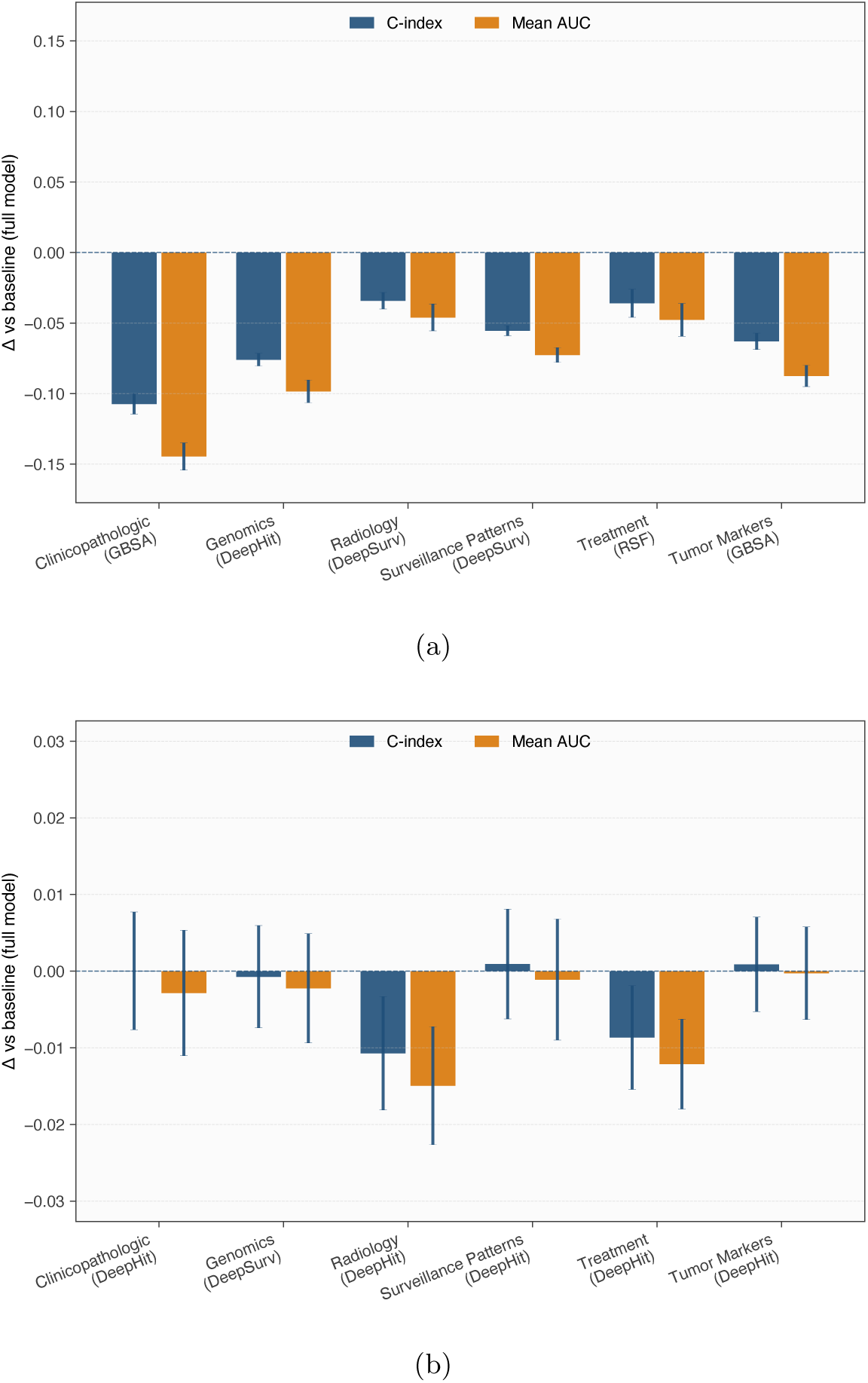
Modality ablations. **a** and **b** Single-modality models and leave-one-modality-out ablations. Bars show the change in C-index and mean AUC relative to the full multimodal model (ablation minus full; negative indicates worse performance), with error bars denoting *±*1 SD across outer folds of nested CV; the best-performing model class for each setting is shown in parentheses.

### Learned risk ranking transfers to external cohorts

We evaluated the five fixed MSK-CHORD GBSA models at DFCI (317 patients, 1,394 regimens) and VICC (152 patients, 695 regimens). The input of this external validation was the original GENIE BPC regimen, and labels the endpoint derived from original PFS-I + Mixed labels, not reconstructed by our mLoT algorithm. C was 0.649 *±* 0.002 at DFCI and 0.634 *±* 0.006 at VICC, with IBS of 0.166 *±* 0.001 and 0.159 *±* 0.001, respectively (Supplementary Table 17). Notably, this performance is achieved despite ECOG, CEA and MMR being unavailable, and CA15–3 was recorded for only 20.8% of DFCI and 3.7% of VICC regimens.

Despite partial transfer of risk ranking, external ICI at 180 days was 11.28 percentage points at DFCI and 12.94 at VICC, with average under-estimation of progression risk (Supplementary Table 25b–c). We therefore evaluated local recalibration to reduce this error in absolute risk estimates, which is a common practice when applied to a new population [24]. We recalibrated with a simple two-parameter logistic update on the log-odds scale at each horizon, while keeping the GBSA models fixed (Methods, “External recalibration”). After the update, ICI at 180 days fell from 11.28 to 0.79 percentage points at DFCI and from 12.94 to 0.71 at VICC (full results in Supplementary Table 25). This suggests substantial improvement achievable with a simple local update, and supports recalibration of absolute probabilities when transferring the model.

### Prognostic stratification despite a front-loaded event distribution

The population Kaplan–Meier (KM) curve computed from the derived PFS labels (red solid line; Fig. 2b) shows frequent early progression and rapid risk-set attrition, with 1,412 (16.1%) mLoTs at risk at 1 year and 552 (6.3%) at 730 d (*≈*2 years). The mean out-of-fold predicted survival curve tracked the population KM curve, with MAE 0.008 and a 3-day difference in RMST at 365 days (Fig. 2b), and their low values suggest predictions capture overall progression dynamics at the population level.

Stratifying the population by predicted 365-day PFS probability yields marked prognostic separation. The high-risk tertile has a KM RMST of 102 d, with only 2% of mLoTs remaining at risk 1 year after line start, compared with 244 d and 35% in the low-risk tertile. MAEs are 0.030, 0.011, and 0.009 in the low-, mid-, and high-risk groups, respectively. The model over-predicts progression in the low-risk tertile, with ΔRMST of *−*10 days overall and *−*18 days in first lines (Figs. 2b and 4e).

**Fig. 4:**
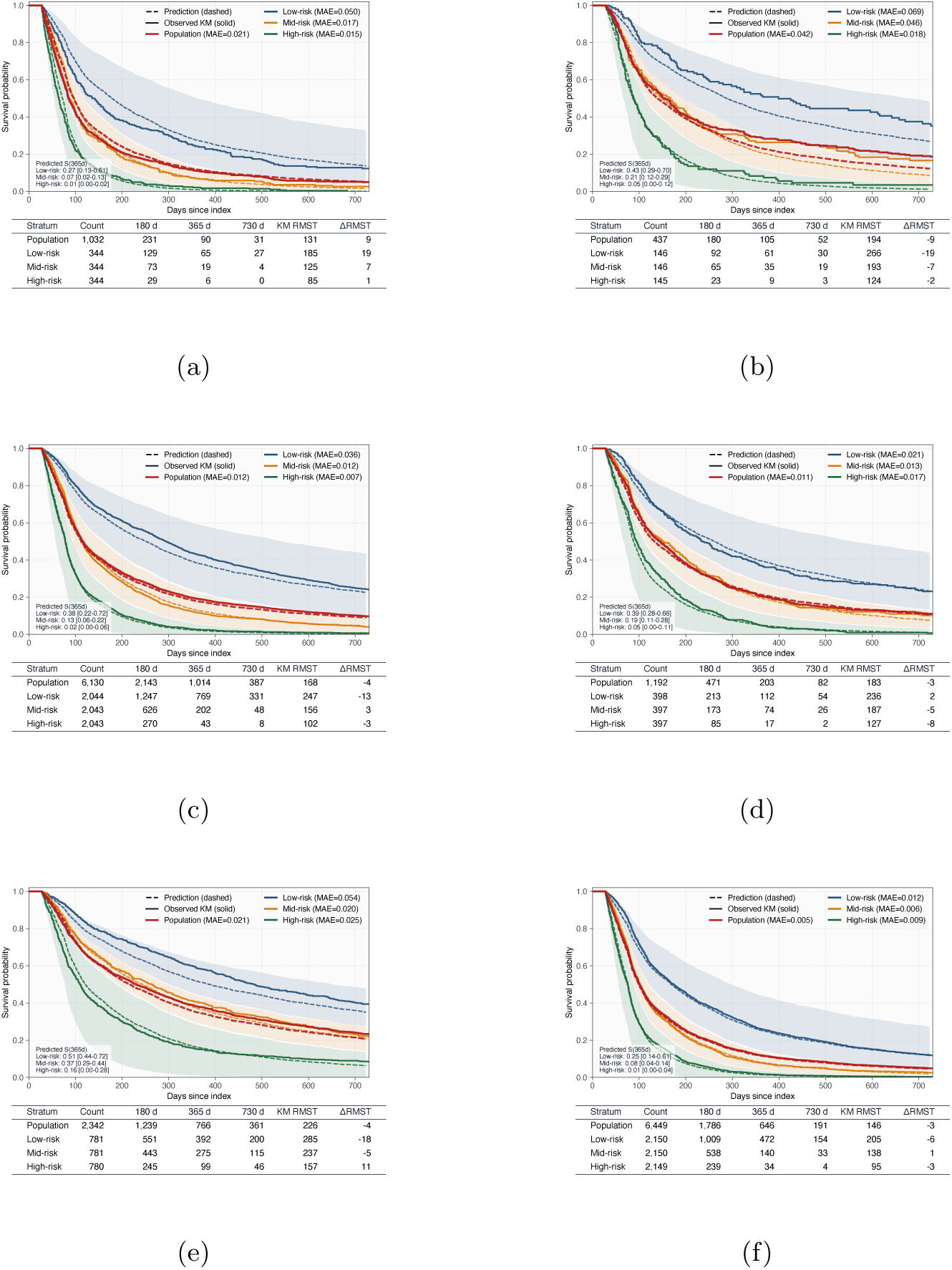
Disease subtype (top/middle) and line-group (bottom) population (red) and tertile-stratified Kaplan–Meier (solid) and mean predicted survival (dashed). **a**: HR*−*/HER2*−*; **b**: HR*−*/HER2+; **c**: HR+/HER2*−*; **d**: HR+/HER2+; **e**: mLoT1; **f**: mLoT2+. Shaded area indicates 95% predicted curves within each tertile. Table shows at-risk counts and RMST summaries. RMST@365: KM RMST with cutoff at 365 days. Δ RMST: predicted RMST - KM RMST. Left legend: range of survival probability at 365 days. Reported 2-year (730 d) summaries are evaluated at the closest available time point on the evaluation grid and labeled as 730 d.

Risk stratification using group-specific thresholds for disease subtypes and line numbers is shown in Fig. 4. We observe substantial differences between high- and low-risk groups within the same cohort, as well as among the same risk group across different cohorts. For example, the RMST differs by 145 days for HR+/HER2*−* and 100 days for TNBC. At the same time, the low-risk tertiles of both HER2+ cohorts have KM-based RMST values above 230 d, compared with 185 d in TNBC; similarly, the low-risk mLoT1 group reaches 285 d versus 205 d in low-risk mLoT2+.

Despite substantial differences in cohort size, with the smallest cohort being HR*−*/HER2+ (437 mLoTs) and the largest HR+/HER2*−* (6,130 mLoTs), agreement between predicted and observed outcomes remains broadly similar. Across risk subgroups, MAE *≤* 0.05 except for the low-risk tertile of HR*−*/HER2+ and the low-risk tertile of mLoT1. *|*ΔRMST*| ≤* 11 d in all non-low-risk cohorts and remains below 20 d in the low-risk tertiles.

### Interpretability analyses highlight disease and care-process signals

Shapley additive explanations (SHAP) [25] for GBSA progression probability output (Fig. 5a) identify the dominant predictive features. At the population level, signals clearly driving higher progression probability include radiology evidence of tumor presence, administration of less frequently used chemotherapy agents, tumor-marker kinetics, ECOG, and TP53 genomic mutation. These features consistently shifted predictions toward higher risk. Lower progression probability drivers include a line being the first metastatic line, absence of genomic sequencing data (likely proxying earlier or less heavily characterized disease in this cohort), and patient age. In line-specific explanations, a low-risk example (Fig. 5b) is associated with absence of disease burden in a recent report, targeted therapy eligibility, and low CA15–3 kinetics; conversely, an mLoT2+ high-risk example (Fig. 5c) is associated with recent liver metastasis, chemotherapy exposure, high CEA and CA15–3 kinetics, and ECOG status. Within subtype and line groups, standardized differences between high- and low-risk tertiles (Cohen’s d) were generally largest for metastatic burden, especially liver involvement, and for first-line and genomic-availability indicators. In mLoT1, recent tumor-marker testing ranked highest (Supplementary Table 5). Group-level attribution shares are contrasted in Supplementary Table 6.

**Fig. 5:**
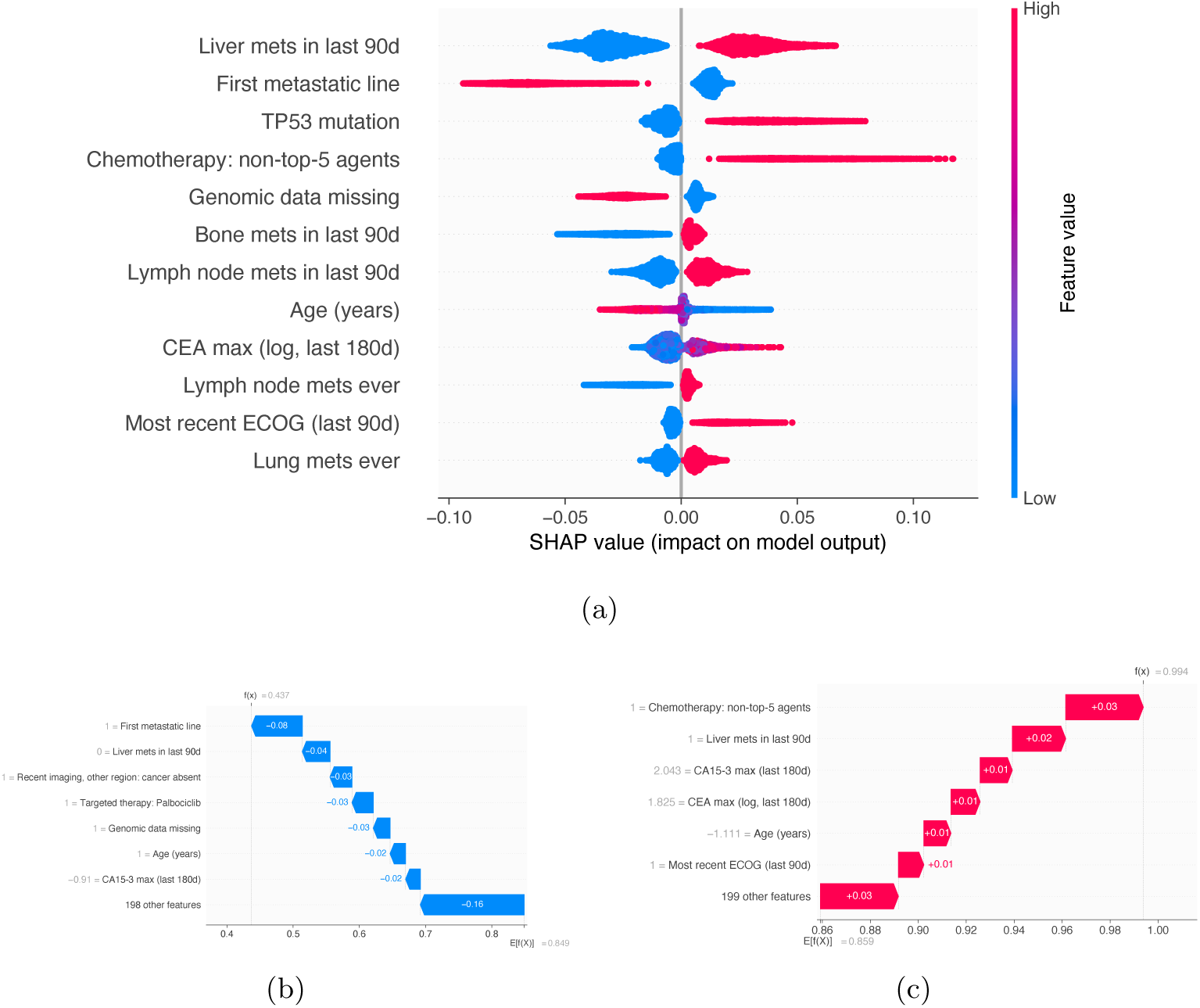
Interpretability analysis for GBSA risk scores. **a** SHAP summary plot for model inputs showing the top 12 contributors to progression probability by 365 days; red indicates higher feature values and positive SHAP values increase progression probability. **b** and **c**: Waterfall plots illustrating line-level explanations for lines from low- and high-risk population tertiles, respectively. *E*[*f* (*x*)] denotes the cohort-mean progression probability and *f* (*x*) denotes the line-specific model output.

Removing surveillance-pattern features had little effect on performance (Fig. 3b), and these features accounted for *≤* 15% of overall SHAP attribution (Supplementary Figure 2). Joint removal of first-line status and all missingness indicators reduced pooled C from 0.6807 to 0.6784. C was 0.6618 within first lines and 0.6568 within later lines, showing that discrimination remained within both line groups (Supplementary Table 24).

## Discussion

In this work, we developed a framework to reconstruct metastatic lines of therapy and model their PFS, especially for later lines, which are less well characterized than their curative and early-line counterparts. In metastatic breast cancer, models trained on MSK-CHORD supported interpretable prognostic stratification, low overall calibration error, and robustness to modality ablation. On external cohorts, reconstructed episodes showed agreement with curated GENIE BPC regimens, and MSK-CHORD models showed partial transferability in risk ranking, and broadly calibrated risk estimates after local recalibration. The evidence supports retrospective prognostic stratification and hypothesis generation, while the potential to aid clinical decision-making and improve patient outcomes remains to be demonstrated.

Disease subtype distribution from the constructed cohort broadly aligns with SEER population statistics [26], with modest TNBC and HER2+ enrichment expected in a metastatic cohort. In mLoT1, regimen backbones align with guideline-consistent practice: chemotherapy-dominant treatment in TNBC, HER2-targeted therapy (often combined with chemotherapy) in HER2+ disease, and endocrine therapy with optional targeted agents in HR+/HER2*−* disease [10]. In mLoT2+, broadly applicable regimen classes (chemo-only, biologic-only, and chemo+biologic) recur across subtypes, consistent with later-line escalation, convergence toward palliative backbones, and increasing use of treatments not restricted to a single biomarker-defined group. At the population level, outcome patterns align with clinical expectations: prognosis is better in earlier lines and poorer in TNBC relative to other subtypes.

Validating our algorithm to the GENIE BPC cohort, we demonstrated that extracted regimens exhibit high regimen agreement with established protocol, with most remaining differences reflecting specific regimen-grouping and endpoint rules (Supplementary Table 19). Our design is intended to preserve long, non-progression lines. Differences arise when BPC records separate regimens after treatment changes without counted progression, and from censoring rules and attribution of progression within the first 28 days.

Central to our approach is the joint modeling across subtypes and lines, motivated by two observations of (especially later) metastatic lines of therapy. First, in real-world EHR records, regimens exhibit substantial heterogeneity beyond a few dominant combinations. Second, regimen classes recur across subtypes, particularly in later lines, creating shared structure that can be leveraged. We show that prognostic modeling benefits from multimodal models that integrate patient-, disease-, and treatment-trajectory features.

Two choices guided feature design. First, we coarsely encode care intensity and surveillance patterns for highly prognostic signals (e.g., radiology and lab measurement) to motivate reliance on biological signals. Consistent with this aim, features designed as surveillance-pattern proxies have minimal impact when ablated and account for only a small fraction of attribution mass (Fig. 3b). Second, we limit model inputs to information recorded before line start, with one exception: drugs received within 28 days of line start. Including treatment administered within 28 days is designed to accommodate treatment scheduling and administration latency (Methods, “mLoT segmentation”). Excluding treatment administered after 28 days is motivated by a leakage pathway, in that including the complete set of drug agents administered within an mLoT improves apparent predictive performance and agreement with observed curves (Supplementary Tables 7–8; Supplementary Figure 4). Presumably, these features implicitly encode post-line information such as drug switches due to intolerance or early signs of progression. They also induce immortal-time-related bias, in that longer PFS mechanically permits more drug administrations. Sensitivity analysis shows that updating the grace period does not substantially impact model performance (Supplementary Tables 11 and 12).

Under this design, the multimodal feature set improves performance more than model class, and also introduces robustness to ablation. The improvement observed within the CoxPH model family supports an incremental contribution from the additional features. In the ablation studies, discriminative power remains stable when certain modalities are missing, within mLoT1 and mLoT2+ groups, and when care-process signals are missing. These findings do not fully establish the absence of care-process reliance, because correlated inputs may retain related information. Adding the number of recorded metastatic-site categories changed C by 0.00054 (95% CI *−*0.00064 to 0.00174; Supplementary Table 26), suggesting that redundancy of data already exists.

When applied to external validation cohort with substantial missingness in input, using labels that are curated with different rules from that of the training set, the model still retains discriminative power. The validation set, which did not require reapplying the evidence enrichment pipeline, reduces a potential source of algorithm-specific validation bias. Minor degradation of model discriminative power (Δ C-index *≤* 0.05) suggests that the model captures transferable prognostic signals. Risk ranking also remained similar under a different endpoint construction, using only the original BPC PFS-I labels, which exclude Mixed imaging assessments (C = 0.643 at DFCI and 0.626 at VICC; Supplementary Table 17). This suggests that the reconstructed labels are sufficiently aligned with independently curated real-world line-level PFS endpoints to support learning prognostic representations that generalize beyond the source cohort. This may be of interest, as different data documentation rules and missingness could be a common theme in medical data.

Across the overall cohort and evaluated subgroups, predicted risk tertiles showed prognostic separation. Internal ICI was 0.91–1.75 percentage points across the evaluated horizons. However, the model overestimated progression in the low-risk tertile, with predicted RMST 10 days below observed overall and 18 days below observed in first lines (Figs. 2b and 4e). After patient-level cross-validated local updates, external ICI was 0.71–1.81 percentage points across the two sites and three horizons. These results support broadly calibrated absolute risk estimates in the evaluated external cohorts after local recalibration. Clinical use still requires evaluation of calibration in the intended population and of the decisions based on those predictions.

Across the overall cohort and within each subtype, GBSA produces risk strata with clear within-group prognostic separation, despite substantial subgroup size imbalance. Line-level attributions describe disease and care-process signals and provide a basis for plausibility checks and hypothesis generation. Several top attributions involve established prognostic domains, including metastatic burden, genomics, and tumor-marker kinetics, and their monotonic directions are biologically sensible (e.g., liver involvement or TP53 alterations should not reduce progression risk). Subtype and initial diagnosis are not among the top contributors, implying that the model relies more on recent disease-burden signals. A plausible explanation is that by mLoT2+, much of the biologic subtype signal has already been “acted upon” through prior regimen selection and earlier outcomes, and the remaining variation in near-term progression risk is increasingly governed by current metastatic burden, marker trajectories, and context- and eligibility-driven treatment choices. Such line-specific attribution shifts describe which signals could be interesting to review at different treatment stages.

Interpretation of some features, however, needs to be put in context. First, the model does not estimate counterfactual treatment effects, and some seeming associations between treatment features and risk are likely confounded. For example, associations such as higher risk with rare chemotherapy or lower risk with targeted/biologic therapy likely reflect confounding by indication and eligibility differences.

Second, missingness in genomic features is associated with lower predicted progression risk. This likely reflects several possible situations, such as patients with genomic data being more likely to have received prior lines, undergone more intensive evaluation, or had more aggressive disease. We retained this signal to represent the absence of genomic data under the line-specific timing rule. In real-world practice, the biological effect of genomic alterations is difficult to separate from the clinical processes that determine whether and when testing occurs, especially given potential time differences between sample acquisition, sequencing, and treatment initiation. Importantly, our ablation results suggest that this proxy does not materially drive performance: removing genomic features caused only negligible performance degradation, while a genomics-only model performed poorly (Fig. 3a). Another example of contextual interpretation is age: higher age is associated with lower predicted risk (Fig. 5a), which is unlikely to be protective biologically. This pattern plausibly reflects cohort composition (older patients enriched for HR-positive and HER2-negative disease; Supplementary Table 1b) and/or more conservative care patterns for older patients. Across the five model families, removing age changed mean outer-fold C by *−*0.00893 to +0.00024 (Supplementary Tables 9 and 10).

This study has several limitations.

First, we model imaging-based PFS under care practices and documentation specific to the source institutions. The modeled population consists of eligible treatment lines under active imaging surveillance at MSK, and generalization to lines without similar follow-up remains uncertain. Prior work has shown that comparability of realworld PFS to trial-based PFS endpoints is limited [27, 28], and that within rwPFS, labels anchored by radiology and clinician can also differ [29]. Our results should be interpreted in light of imaging-based PFS, similar to GENIE BPC’s imaging-based labels, rather than as directly interchangeable with clinician-assessed or trial-based PFS. We compare extracted HR+/HER2*−* PFS on letrozole plus palbociclib, and letrozole against a real-world PFS study [30], since it is the most directly comparable. The study reported weighted clinician-assessed real-world PFS of 20.0 and 11.9 months, with some weighting to balance the study’s treatment group. Among first HR+/HER2*−* metastatic lines, median PFS was 14.9 months with letrozole plus palbociclib and 12.6 months with letrozole alone in our cohort. These medians were 454 and 382 days, compared with 289 and 254 days, respectively, for the same named regimens using endpoint derived from original labels from the GENIE BPC cohort (Supplementary Table 20). GENIE BPC provides a curated external reference for treatment regimens and imaging-based outcomes. Differences in regimen grouping, progression attribution, and censoring conventions should be considered when interpreting reconstruction agreement (Supplementary Table 19).

A line can contain several imaging reports, yet the impact of report-level NLP false positives on line-level PFS remains hard to quantify without access to original reports. Labels are assigned from a report’s impression section, but it is not explicitly stated whether reports share a common baseline [17]. Therefore, reports within the same line may not all refer to the patient status at the treatment start, and successive reports may refer to different patient statuses in time. Under rules requiring no subsequent non-progression report within 30, 60 or 90 days, first-line C remained 0.661–0.664, while median PFS increased from 231 days to 274, 311 and 378 days (Supplementary Table 21). Furthermore, independent review in the original MSK-CHORD study attributed 42 of 50 apparent false-positive progression calls to curation errors instead of model error [17]. Although the exact impact remains unquantified, the above two results suggest that annotation accuracy could be reliable, and our central conclusion may not be heavily impacted by the NLP label error rate.

Informative censoring remains a limitation in real-world PFS analyses. In our setting, informative censoring may occur when an mLoT advances to a subsequent line without a qualifying progression label, and is instead censored at the last recorded nonprogression assessment. Although the non-progression status was documented, if the eventual treatment change was related to undocumented disease worsening, outcomes for these lines may differ from those remaining under observation, potentially biasing PFS estimates [31, 32]. This population that could be impacted is bounded above by 674/8,791 mLoTs (7.7%). Because this mechanism can arise only for lines assigned non-progression censoring, and because administrative censoring at 2 years is noninformative by design, only mLoTs censored before the 2-year cutoff may be affected by this bias. Moreover, some of these lines likely reflect non-progression-related treatment changes, such as toxicity, patient preference, or planned escalation, which would not constitute informative censoring. Addressing this limitation will require higher-resolution routine-care data and future work on richer data capture and modeling approaches.

A related selection effect arises from the QC rules. For 525 lines (4.7% of the 11,085 eligible lines) in which at least one treatment class first appeared after day 28, the chemotherapy plus endocrine exclusion was not independent of outcome (discussed in detail in Methods, “PFS labeling and censoring”). As a result, the modeled cohort under-represents favorable outcomes in HR-positive lines starting on chemotherapy. We report the original cohort alongside refits that restore these lines. The refits changed overall discrimination little and reduced mean under-prediction in these subgroups, but did not improve external calibration (Supplementary Table 28).

Finally, the model uses treatment exposure recorded within the initial 28-day window and is not established as a prospective day-zero predictor. This constraint follows from the leakage-prevention design discussed above and from the model not updating risk dynamically within a line. Therefore, analyses are strictly retrospective, and prospective clinical validation is needed before the model is used to inform patient care.

There are many directions that future work could pursue. First, mLoT segmentation methods that integrate structured therapy timing with radiology and clinical signals, as well as holistically review multiple radiology labels throughout the trajectory, could reduce reliance on heuristics and improve boundary accuracy. Scalable weak supervision or clinician-in-the-loop audits could further provide training signal and ongoing quality control. Second, adapting dynamic time-series models that update risk within a line and across successive mLoTs at predefined, leakage-safe update times could considerably improve model performance. Third, higher-fidelity inputs (raw imaging, longitudinal labs/vitals, free-text notes, and richer molecular profiling) could improve prognostic resolution, especially in later-line heterogeneous disease. These extensions could broaden applicability beyond curated cohorts and improve prognostic stratification, subject to further validation across diseases and institutions.

## Methods

### Cohort construction

We included MSK-CHORD patients with breast cancer as the single primary malignancy who had palliative metastatic treatment records and radiology-derived progression assessments. Metastatic onset was approximated as the earliest of (i) the first mention of a distant metastatic site in radiology reports (excluding regional lymph nodes and nonspecific sites) or (ii) the diagnosis date for de novo stage IV disease.

### mLoT segmentation

We separately perform mLoT segmentation and PFS labeling to allow for modularity and anchor derived pairs to recorded radiology-based progression assessments. The segmentation algorithm uses systemic treatment records and NLP-annotated radiology progression assessments. For each patient, mLoT1 begins at the first systemic therapy start date after metastatic onset and advances according to the following rules, reflecting clinical conventions and oncologist input.

Importantly, the rules below are used only for mLoT segmentation and never directly label PFS events.

- An mLoT advances after an NLP-labelled radiology progression event that occurs at least 28 days after the current line start date;
- An mLoT advances after a new systemic therapy drug (not the preceding drug) that starts at least 1 year after the current line start date;
- An mLoT does not advance after local treatment changes (e.g., bone-directed agents, surgery, radiation therapy);
- The next mLoT begins at the start date of the first subsequent systemic therapy.

All treatments administered between the start and end of an mLoT are considered part of the mLoT for downstream filtering. An exception applies when a new systemic treatment is initiated within 28 days before the recorded progression that ends the current mLoT. In that case, it is assigned to start a new mLoT to avoid attributing progression to a regimen with insufficient exposure or radiology-assessment latency.

The 28-day threshold is a pragmatic compromise based on clinician input: it allows routine scheduling and assessment latency around line start while avoiding attribution of an observed progression to a regimen with essentially no exposure. Comparable operational conventions for line-of-therapy analyses have also been used in other oncology real-world studies [13, 14, 33, 34]. Sensitivity analyses using narrower and wider line-start treatment windows (14 and 42 days) yielded nearly unchanged discrimination and calibration metrics, supporting robustness of the treatment-window choice rather than dependence on the specific 28-day cutoff (Supplementary Tables 11 and 12).

### PFS labeling and censoring

For survival analysis, we label each segmented mLoT with a PFS(event, time) pair. The event label is binary, indicating whether the line ended with progression or death, or was censored. The PFS time is defined as the number of days from line start to the label-defining event, and is not the time between the start of two consecutive mLoTs. We use the following rule set to assign PFS labels:

- Progression event: an mLoT is labeled as a progression event if it is terminated by a qualifying NLP-labelled radiology progression event. The time between line start and the first such progression event is the PFS time;
- Death event: for the final mLoT, death without a prior documented progression is also counted as a progression event. The time between line start and death is the PFS time;
- Right censoring: if neither of the above occurs, the line is right-censored at the time of the last radiology-confirmed non-progression assessment. PFS time is the number of days from line start to that last non-progression assessment;
- Exclusion: lines without any follow-up radiology assessment or death event are not assigned PFS labels and are excluded from modeling.

Radiology-confirmed progression was determined using the progression-status labels provided by MSK-CHORD. We did not develop or re-infer these NLP labels. We treated them as an upstream validated endpoint component, as the original study reported an AUC of 0.97 with precision and recall of 0.90 for progression-status identification [17] against manually curated labels.

We additionally apply a pragmatic rule set to further strengthen mLoT regimen and PFS label fidelity. Constructed mLoT-PFS pairs are excluded if:

- No radiology assessment or death event occurred during the mLoT;
- No radiology assessment occurred within 90 days prior to the start of the mLoT;
- The assigned PFS duration is shorter than 28 days;
- Both chemotherapy and endocrine therapy were administered within the mLoT;
- HR-negative patients received endocrine therapy.

Among 2,294 QC exclusions, 2,114 involve chemotherapy and endocrine therapy within the same mLoT, and 261 involve endocrine therapy in HR-negative disease. Exclusion of lines containing both chemotherapy and endocrine therapy presents an interesting “trilemma”. Encoding both classes as simultaneous inputs may lead the model to treat implausible co-prescriptions as normal patterns of care. Encoding the eventual sequence at line start introduces immortal-time bias, since only patients with long PFS go on to receive the later class. Encoding the first class and masking the later class attributes the outcome of the whole line to the drugs given at line start. We used the filter to limit this ambiguity in the treatment representation. Our BPC comparison supports this concern: in all 127 flagged late-start episodes, the later class belonged to a separate BPC regimen (Supplementary Table 28f). However, exclusion has its own cost: for later treatment additions, it selects on remaining progression-free until that treatment is recorded. In 1,589 of the 2,114 flagged lines both classes were recorded within the first 28 days; in 525 (4.7% of the 11,085 eligible lines) the second class was added later. Refitting with the 505 restorable lines, with later agents masked, did not change risk ranking (paired change in C *−*0.0008, 95% CI *−*0.0017 to +0.0001). It reduced the under-prediction of one-year PFS in HR-positive first lines starting on chemotherapy (17.0 to 10.5 points in HR+/HER2+; 9.7 to 3.4 in HR+/HER2*−*) but did not improve external calibration (Supplementary Table 28c–d). Evaluating the saved GBSA models on all 11,085 eligible lines with the 2,294 QC-excluded lines restored left pooled concordance essentially unchanged (C 0.6807 versus 0.6815; C 0.6953 within the QC-excluded lines alone; Supplementary Table 15). The 90-day pre-line radiology requirement provides a documented baseline disease-state assessment and defines the target population as eligible lines under active imaging surveillance within the institution. This reduces bias from incomplete capture (e.g., outside-system care) that could distort PFS and induce model reliance on non-biological proxies (Supplementary Figure 3). Together, these rule-based filters prevent contradictions with standard-of-care patterns and limit spurious correlations between documentation intensity, subtype labels, and hazard.

We also apply administrative censoring at 730 days (2 years) to mitigate the influence of long-term survivors with limited follow-up and to focus on clinically relevant risk stratification. In the final analytic cohort, 1,226 of 8,791 mLoTs are censored, with 674 lines censored after a documented non-progression assessment and 552 administratively censored at 730 days (Fig. 1b). This design separates mLoT advancement from PFS labeling and makes the progression and censoring rules explicit.

For the first-line endpoint sensitivities, we evaluated rules that required no subsequent non-progression report within 30, 60, or 90 days after a positive report (Supplementary Table 21). For the imaging-eligibility sensitivity, we extended the pre-line imaging requirement from 90 to 180 days and evaluated the saved GBSA models. Performance remained essentially unchanged (pooled C 0.6807 versus 0.6828; Supplementary Table 27).

### Feature extraction

Features are extracted and engineered from the MSK-CHORD dataset and are grouped into five modalities: clinicopathologic, radiology, tumor markers, treatment regimen, and genomics (Supplementary Note 1 Feature variables). The model has 205 inputs: 36 treatment, 52 genomic, 36 clinicopathologic, 36 tumor-marker, and 45 radiology inputs (Supplementary Table 18). The feature-extraction pipeline is available in the study code repository. Clinicopathologic features summarize baseline demographics and disease characteristics. Radiology features are derived from radiology reports within 90-day windows prior to line start, capturing tumor presence in specific organs and cancer presence from imaging of different body regions (several regions share the same report outcomes). Tumor-marker features summarize CA15–3 and CEA kinetics using various log-transformed aggregation statistics. Treatment features encode drug agents and local therapies administered within 28 days of line start, using the top five plus an “other” category for each systemic treatment. Genomic features encode somatic mutations and copy-number aberrations in a curated gene list. Genomic features are masked for pre-sequencing lines, with an additional missingness flag for lines that precede sequencing. Under this time-restricted encoding, genomic features are masked for 3,010 of 8,791 modeled mLoTs, spanning 1,805 of 2,881 patients. Additionally, we include a binary flag to indicate whether a line is the first metastatic line, to account for the established prior that treatment benefit tends to diminish across successive lines. Numeric features were centered by the median and scaled by the interquartile range estimated from the training fold, and the same transformation was applied to validation and test data. Missing numeric measurements were then set to zero, which represents the training-fold median. Missing binary indicators were set to zero or encoded with an unknown category, as specified for each feature. Missingness flags, last day of observation, and cancer-presence report status by imaged body region are additionally considered as a “surveillance patterns” modality.

Two design choices specifically mitigate information leakage and non-biological shortcuts. First, drug agents initiated *>* 28 days after line start are excluded from model inputs. Since mLoTs are constructed retrospectively, encoding the full within-line treatment record risks introducing bias. Longer PFS mechanically permits more post-initiation modifications, such as toxicity switches or drug add-on. As a result, the model may learn a shortcut to associate more drug agents received with longer PFS. The masking is justified by the following observations. mLoTs with agents initiated *>* 28 days after line start exhibit substantially different PFS distributions from those without late-start agents (Supplementary Figure 4, dashed lines). Models trained without late-start agent masking produced mean predicted curves closer to the corresponding KM estimates and higher apparent discrimination (Supplementary Tables 7 and 8; Supplementary Figure 4).

Second, we applied conservative feature engineering to mitigate potential reliance on surveillance patterns. Because progression is observed through clinical surveillance, models can learn care-process proxies rather than underlying disease risk. For example, given diminishing benefit across successive mLoTs, the model may learn to associate heavier prior treatment with shorter subsequent PFS. We therefore omit explicit treatment trajectories, under the assumption that the current regimen already reflects clinician synthesis of prior response and tolerability. We also suppress finegrained surveillance by omitting measurement counts and timestamps in biomarker kinetics and radiology screening, except for the most recent day of observation and value. Surveillance pattern variables are evaluated in addition to clinical modalities in ablation experiments.

### Model selection and training

We benchmarked the following survival models that span varying complexity, adherence to the PH assumption, and interpretability:

- (PH, linear) CoxPH [21, 22].
- (PH, tree-based) Gradient-boosted survival model with CoxPH loss (GBSA).
- (Non-PH, tree-based) Random survival forest (RSF) [35].
- (PH, deep learning) DeepSurv [36].
- (Non-PH, deep learning) DeepHit [37].

Code for model implementations and evaluation metrics is adapted from the framework and Python notebooks in [38].

We performed nested cross-validation (nested CV) with 5 outer folds for evaluation and 3 inner folds for hyperparameter selection. This provides an unbiased estimate of held-out performance under hyperparameter exploration and model selection. For GBSA, RSF, DeepSurv, and DeepHit, we evaluated five preselected configurations (Supplementary Note 2 Model parameters) spanning low-, medium-, and high-capacity models with different regularization strengths. For CoxPH, we tuned the L2 regularization strength *α* over 30 log-spaced values between 10*^−^*^1^ and 10^3^. Best-performing configuration candidates in each outer fold were those with average inner-fold C-index within 0.003 of the best-performing configuration. The final configuration is selected in favor of lower-complexity and more strongly regularized configurations. CV splits were stratified by HR/HER2 subtype and whether a patient had at least one mLoT censored at 2 years. All mLoTs from a given patient were assigned to the same split to prevent leakage. Detailed candidate grids are provided in Supplementary Note 2 Model parameters, and selected outer-fold configurations are summarized in Supplementary Table 14. Ablation experiments use the selected best configuration, restricted to configurations selected at least once in the full-modality nested CV to reduce computational burden.

The clinical Cox reference used first-line status, HR and HER2 status, the most recent ECOG value, liver involvement in the preceding 90 days, and the most recent CA15–3 value within 180 days before line start, transformed as log(1 + *x*), with *α* = 1 (Supplementary Table 23). Additional GBSA analyses jointly removed first-line status and all missingness indicators, or added the number of recorded metastatic-site categories in the preceding 90 days (Supplementary Tables 24 and 26).

### Evaluation

We harmonized each model’s output to a survival probability curve on a common time grid over 2 years. Global discrimination metrics are Antolini’s C [39] and cumulative/dynamic AUC (cdAUC) [40]. Antolini’s C considers all comparable pairs of mLoTs over follow-up and reports the fraction for which the line with the shorter observed PFS receives higher predicted risk. cdAUC is computed at each time *t* by forming the time-specific risk set, pairing individuals who experience an event prior to time *t* with those who remain event-free past *t*, and estimating the probability that the model assigns higher risk to the former; inverse-probability weights correct for censoring. An average over the evaluation time range yields the reported single overall discrimination score. For time-dependent evaluation, we report IPCW C-index [41], cdAUC, and Brier score [42] at 30-day increments. Inverse-probability-of-censoring weights were estimated with the Kaplan–Meier method in the population being evaluated: the MSK-CHORD training folds for internal evaluation and each external site’s own regimens for external evaluation. To prevent degenerate inverse-probability-of-censoring weights at the administrative-censoring boundary, any PFS event occurring exactly at 730 d is shifted to 729.999 d (i.e., 730*−*0.001). This negligible adjustment avoids censoring-survival estimates reaching 0 at the evaluation horizon and stabilizes IPCW-based metrics. For the same reason, IPCW-based metrics at the 2-year horizon are evaluated at 730*−ε* rather than exactly 730 d. When a nominal horizon (e.g., 730 d) is not exactly represented on a model’s evaluation grid, we evaluate at the closest available time point and report it at the nominal horizon.

Pooled out-of-fold estimates were reported with percentile 95% confidence intervals from 1,000 patient-level bootstrap resamples, retaining all lines of each sampled patient. Model differences used paired patient draws. Predictions remained fixed, so these intervals describe evaluation-sample uncertainty and do not include uncertainty from model training.

Calibration was evaluated at 180, 365, and 730 days using IPCW, with ICI calculated from a three-knot natural cubic-spline calibration curve and reported in percentage points. Calibration-in-the-large is the intercept from an IPCW logistic model with the coefficient of predicted log-odds fixed at one; calibration slope is estimated with a free intercept. Their ideal values are zero and one, respectively; positive calibration-in-the-large indicates average under-estimation of progression risk, and a slope above one indicates compressed predictions. ICI, E50, and E90 are the mean, median, and 90th percentile of absolute calibration error, respectively. Internal probabilities are out-of-fold, whereas external probabilities are averaged across the five fixed fold models. Calibration intervals use 1,000 patient resamples, retaining all lines of each sampled patient; censoring estimates and calibration models are refitted, while prediction models and spline knots remain fixed. Additional calibration measures—calibration-in-the-large, calibration slope, E50, and E90—are reported in Supplementary Table 25. The completed TRIPOD+AI checklist is provided in Supplementary Note 3.

### External recalibration

At each external site and at 180, 365, and 730 days, we fitted an IPCW-weighted logistic model to estimate an intercept and slope for the log-odds of the mean predicted progression probability from the five fixed GBSA models. The updated log-odds equalled the intercept plus the slope multiplied by the original log-odds. We evaluated the update using five patient-level folds within each site; each regimen received an update fitted on patients in the other four folds. Recalibration intervals used 1,000 patient-level bootstrap resamples with the updated probabilities held fixed; the update was not refitted within each resample. For the pooled external ICI summary, we combined the DFCI and VICC regimens and fitted the same calibration diagnostic with pooled censoring weights to the original or site-specific cross-validated updated probabilities. This pooled estimate is distinct from the site-specific results. Continuous calibration curves and observed versus predicted survival within external risk tertiles are shown in Supplementary Figures 5–6.

### Reconstruction comparison on GENIE BPC

We applied the mLoT segmentation and PFS-labeling rules to curated GENIE BPC treatment and imaging records at DFCI, VICC, and MSK. A reconstructed episode and a BPC regimen were matched within the same patient if they had identical drug sets after drug-name normalization and removal of GnRH agonists (since GENIE BPC counts these as separate regimens), with start dates at most 14 days apart. We reported regimen matching among retained reconstructed episodes and event-indicator and exact-duration agreement among matched pairs. Listed grouping and endpoint-rule differences were reported separately from exact agreement (Supplementary Table 19).

### External validation on the AACR Project GENIE BPC cohort

In external validation, the five GBSA models trained on the MSK-CHORD outer folds were applied unchanged. We used curated GENIE BPC metastatic regimen boundaries, which follow BPC grouping rules and are not equivalent to reconstructed MSK-CHORD mLoTs. We derived PFS-I + Mixed from curated imaging assessments, counting the first Progressing or Mixed report after regimen start, or death, as an event; censoring follows the BPC next-regimen and last-contact rules [20]. We included Mixed assessments as progression in BPC PFS-I to match the positive class of the MSK-CHORD radiology labels. We retained the original regimen and drug eligibility rules, reapplied the minimum 28-day PFS duration to the revised endpoint, and administratively censored at 730 days. The main analysis uses this endpoint; Supplementary Table 17 also retains the released PFS-I analysis for comparison.

Not all model inputs are available in GENIE BPC. ECOG performance status, CEA, MMR status, and surgery and radiation are not released; these inputs were set to missing for every regimen, as for an MSK-CHORD line without that measurement. CA15–3 was recorded for 20.8% of DFCI and 3.7% of VICC regimens, and PD-L1 status was unknown for nearly all regimens. Radiology-report mentions of tumor sites are not released; the metastatic-site inputs were instead derived from the curated dates of first distant metastasis at each site. All other inputs were built from curated BPC fields.

### Risk stratification and Kaplan–Meier summaries

Risk stratification and interpretability analyses are performed on out-of-fold predictions from the primary model (GBSA). mLoTs are stratified into low-, mid-, and high-risk tertiles using the predicted 1-year progression probability, and we compare the mean predicted survival curve to the Kaplan–Meier estimate within each stratum. Risk tertile cutoffs and SHAP contrasts were calculated at line level. For proportional-hazards models, the risk ranking is time-invariant and thus the same tertile cutoffs apply across time horizons. We summarize the discrepancy using the mean absolute error (MAE) between predicted and observed survival curves over time; a low MAE means the predicted survival probabilities differ from the corresponding Kaplan–Meier estimate by only a small amount on average. We additionally report RMST at 365 days, which informs the expected time free of progression or death within the first year of treatment.

### SHAP contributions

We interpret GBSA predictions at the feature-group level using SHAP values computed from SHAP’s PermutationExplainer on the predicted PFS probability *at* 365 days. Although models trained on different folds may learn different baseline hazards and risk-score calibrations, the predicted PFS probability at a fixed time point is directly comparable across folds. To intuitively correlate higher SHAP output values with higher progression risk, we report SHAP contributions for 1 *−* PFS(365) (i.e., progression probability *by* 365 days).

### Input-feature value contrasts

Let *x_ij_* denote the value of input feature *j* for line *i* under the feature windows defined above. To report subtype(or mLoT-) specific contrasts relative to the cohort baseline, we re-center each input feature within cohort *G*,

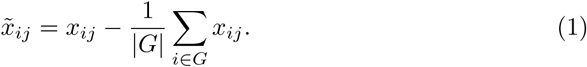

We then quantify separation between risk strata using Cohen’s *d* computed on *x̃_ij_* within cohort *G*:

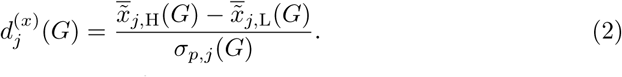

Positive *d*^(*x*)^_*j*_ (*G*) indicates higher mean feature values in the high-risk tertile, while negative values indicate higher mean feature values in the low-risk tertile. We apply this same input-feature value contrast within each HR/HER2 subtype and within each mLoT stratum (mLoT1 and mLoT2+).

### SHAP-based contrasts at the group level

Let *ϕ_ij_* denote the SHAP attribution for feature *j* on line *i* for the prediction 1 *−* PFS*_i_*(365). We additionally map model inputs into clinically motivated feature groups (Supplementary Table 16) and summarize each group’s contribution using a per-line relative-importance (share) score,

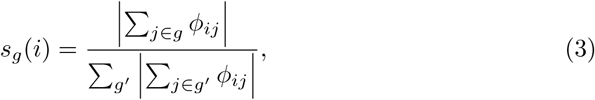

where *g* denotes a feature group and the denominator sums absolute SHAP magnitudes across groups. This normalization yields the fraction of total absolute attribution assigned to group *g* for line *i* and avoids sign cancellation across groups. We compare these group shares directly between risk strata within cohort *G* (without re-centering),

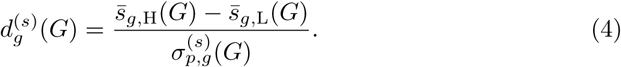

Positive *d*^(*s*)^_*g*_ (*G*) indicates that group *g* accounts for a larger share of total absolute attribution in the high-risk tertile; negative values indicate a larger share in the lowrisk tertile.

### Cohen’s ***d*** and pooled variance

For any scalar quantity *z*(*i*) compared between H and L within cohort *G* (e.g., *x̃_ij_* or *s_g_*(*i*)), we compute Cohen’s *d* as

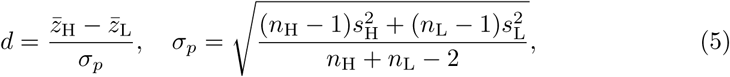

where *z̄*_H_ and *z̄*_L_ are means in the high- and low-risk tertiles, *s*^2^ and *s*^2^ are their sample variances, and *n*_H_, *n*_L_ are the tertile sizes. Throughout, these attributions and contrasts are descriptive and non-causal in this observational setting.

## Data Availability

The dataset analyzed during the current study is the MSK-CHORD dataset [17], released by Memorial Sloan Kettering Cancer Center. It is available via cBioPortal under study identifier msk chord 2024: https://www.cbioportal.org/ study/summary?id=msk chord 2024. AACR GENIE BPC data access instructions can be found in https://www.aacr.org/professionals/research/aacr-project-genie/ aacr-project-genie-data/.

## Code Availability

Code for data generation, modeling, and analysis is available under https://github.com/uzh-dqbm-cmi/mbc-pfs.

## Acknowledgements

The authors would like to acknowledge the American Association for Cancer Research and its financial and material support in the development of the AACR Project GENIE registry, as well as members of the consortium for their commitment to data sharing. Interpretations are the responsibility of study authors.

## Author Contributions

X.Z., B.F., Z.B. and M.K. conceived the study. B.F. and M.K. supervised the study. B.F. advised on the machine learning modeling and feature set design. Z.B. and A.W. advised on algorithm design, medical content, and clinical interpretation. X.Z. implemented the mLoT extraction algorithm, machine learning modeling, and analysis pipeline. T.N. ran modeling experiments with optimized model parameters, performed exploratory data analysis, and refined manuscript figures. X.Z. wrote the manuscript with feedback and guidance from all co-authors, who all contributed to the interpretation of results and refinement of the manuscript.

## References

[1] Cardoso, F. et al. 4th ESO–ESMO International Consensus Guidelines for Advanced Breast Cancer (ABC 4). Annals of Oncology 29, 1634–1657 (2018). URL https://pmc.ncbi.nlm.nih.gov/articles/PMC7360146/.

[2] Fietz, T. et al. Palliative systemic therapy and overall survival of 1,395 patients with advanced breast cancer - Results from the prospective German TMK cohort study. Breast 34, 122–130 (2017).

[3] U.S. Food and Drug Administration. Clinical trial endpoints for the approval of cancer drugs and biologics. Guidance for Industry, Silver Spring, MD (2018). URL https://www.fda.gov/regulatory-information/search-fda-guidance-documents/clinical-trial-endpoints-approval-cancer-drugs-and-biologics. Docket FDA-2005-D-0225.

[4] Cardoso, F. et al. International Guidelines for Management of Metastatic Breast Cancer: Combination vs Sequential Single-Agent Chemotherapy. JNCI: Journal of the National Cancer Institute 101, 1174–1181 (2009). URL 10.1093/jnci/djp235.

[5] Varma, G. et al. A Deep Learning-Enabled Workflow to Estimate Real-World Progression-Free Survival in Patients With Metastatic Breast Cancer: Study Using Deidentified Electronic Health Records. JMIR cancer 11, e64697 (2025).

[6] Merzhevich, T. et al. Machine Learning Predictions of Overall and Progression-Free Survival in Advanced Breast Cancer. Artificial Intelligence in Medicine (AIME 2025), Lecture Notes in Computer Science 267–271 (2025). URL 10.1007/978-3-031-95841-050.

[7] Cabel, L. et al. Outcome beyond third-line chemotherapy for metastatic triplenegative breast cancer in the French ESME program. Breast 56, 18–25 (2021).

[8] Cottu, P. et al. Attrition between lines of therapy and real-world outcomes of patients with HER2-positive metastatic breast cancer in Europe: a cohort study leveraging electronic medical records. Breast Cancer Research and Treatment 209, 419–430 (2025). URL 10.1007/s10549-024-07506-4.

[9] Basmadjian, R. B. et al. Retrospective database analysis to assess treatment patterns, clinical outcomes, and healthcare resource utilization in metastatic or recurrent HER2-positive breast cancer in Alberta. Cancer Treatment and Research Communications 45, 101021 (2025).

[10] Gennari, A. et al. ESMO Clinical Practice Guideline for the diagnosis, staging and treatment of patients with metastatic breast cancer. Annals of Oncology: Official Journal of the European Society for Medical Oncology 32, 1475–1495 (2021).

[11] Saini, K. S. & Twelves, C. Determining lines of therapy in patients with solid cancers: a proposed new systematic and comprehensive framework. British Journal of Cancer 125, 155–163 (2021). URL 10.1038/s41416-021-01319-8.

[12] Meng, W. et al. Temporal phenotyping by mining healthcare data to derive lines of therapy for cancer. Journal of Biomedical Informatics 100, 103335 (2019).

[13] Meng, W., et al. An Automated Line-of-Therapy Algorithm for Adults With Metastatic Non–Small Cell Lung Cancer: Validation Study Using Blinded Manual Chart Review. JMIR Med Inform 9, e29017 (2021). URL http://www.ncbi.nlm.nih.gov/pubmed/34636730.

[14] Grady, C. B. et al. Determining Line of Therapy from Real-World Data in Non-Small Cell Lung Cancer. Pharmacoepidemiology and Drug Safety 33, e70049 (2024).

[15] Kehl, K. L. et al. Assessment of Deep Natural Language Processing in Ascertaining Oncologic Outcomes From Radiology Reports. JAMA oncology 5, 1421–1429 (2019).

[16] Kehl, K. L. et al. Natural Language Processing to Ascertain Cancer Outcomes From Medical Oncologist Notes. JCO clinical cancer informatics 4, 680–690 (2020).

[17] Jee, J. et al. Automated real-world data integration improves cancer outcome prediction. Nature 636, 728–736 (2024). URL 10.1038/s41586-024-08167-5.

[18] AACR Project GENIE Consortium. AACR Project GENIE: Powering Precision Medicine through an International Consortium. Cancer Discovery 7, 818–831 (2017).

[19] Lavery, J. A., et al. A scalable quality assurance process for curating oncology electronic health records: The project genie biopharma collaborative approach. JCO Clinical Cancer Informatics e2100105 (2022). URL https://ascopubs.org/doi/abs/10.1200/CCI.21.00105. PMID: 35192403.

[20] GENIE BPC Statistical Coordinating Center, Memorial Sloan Kettering Cancer Center. GENIE BPC Analytic Data Guide: BrCa v1.0-public. American Association for Cancer Research (2025). URL https://www.aacr.org/wp-content/uploads/2025/12/GENIE-BPC-BrCav1.0-public-Analytic-Data-Guide.pdf. Accessed 11 Oct 2026.

[21] Cox, D. R. Regression models and life-tables. Journal of the Royal Statistical Society. Series B (Methodological) 34, 187–220 (1972). URL http://www.jstor.org/stable/2985181.

[22] Pölsterl, S. scikit-survival: A Library for Time-to-Event Analysis Built on Top of scikit-learn. Journal of Machine Learning Research 21, 1–6 (2020). URL http://jmlr.org/papers/v21/20-729.html.

[23] Austin, P. C. & Steyerberg, E. W. The Integrated Calibration Index (ICI) and related metrics for quantifying the calibration of logistic regression models. Statistics in Medicine 38, 4051–4065 (2019).

[24] Van Calster, B. et al. Calibration: the Achilles heel of predictive analytics. BMC Medicine 17, 230 (2019). URL 10.1186/s12916-019-1466-7.

[25] Lundberg, S. & Lee, S.-I. A unified approach to interpreting model predictions (2017). URL https://arxiv.org/abs/1705.07874.arXiv:1705.07874.

[26] National Cancer Institute (NCI), Surveillance, Epidemiology, and End Results (SEER) Program. Cancer stat facts: Female breast cancer subtypes. https://seer.cancer.gov/statfacts/html/breast-subtypes.html (2026). Accessed 11 Jan 2026.

[27] Panageas, K. S., Ben-Porat, L., Dickler, M. N., Chapman, P. B. & Schrag, D. When you look matters: the effect of assessment schedule on progression-free survival. Journal of the National Cancer Institute 99, 428–432 (2007).

[28] Ackerman, B. et al. Measurement error and bias in real-world oncology endpoints when constructing external control arms. Frontiers in Drug Safety and Regulation **Volume** 4 - 2024 (2024). URL https://www.frontiersin.org/journals/drug-safety-and-regulation/articles/10.3389/fdsfr.2024.1423493.

[29] Griffith, S. D. et al. Generating Real-World Tumor Burden Endpoints from Electronic Health Record Data: Comparison of RECIST, Radiology-Anchored, and Clinician-Anchored Approaches for Abstracting Real-World Progression in Non-Small Cell Lung Cancer. Advances in Therapy 36, 2122–2136 (2019).

[30] DeMichele, A. et al. Comparative effectiveness of first-line palbociclib plus letrozole versus letrozole alone for HR+/HER2-metastatic breast cancer in US real-world clinical practice. Breast Cancer Research 23, 37 (2021). URL 10.1186/s13058-021-01409-8.

[31] Fleming, T. R., Rothmann, M. D. & Lu, H. L. Issues in using progression-free survival when evaluating oncology products. Journal of Clinical Oncology 27, 2874–2880 (2009). URL https://ascopubs.org/doi/abs/10.1200/JCO.2008.20.4107. PMID: 19414672.

[32] Campigotto, F. & Weller, E. Impact of informative censoring on the Kaplan-Meier estimate of progression-free survival in phase II clinical trials. Journal of Clinical Oncology: Official Journal of the American Society of Clinical Oncology 32, 3068–3074 (2014).

[33] Bains, S., Kalsekar, A., Amiri, K. I. & Weiss, J. Real-World Treatment Patterns and Outcomes Among Patients With Metastatic NSCLC Previously Treated With Programmed Cell Death Protein-1/Programmed Death-Ligand 1 Inhibitors. JTO Clinical and Research Reports 3 (2022). URL 10.1016/j.jtocrr.2021.100275.

[34] Barzi, A. et al. Real-World Dosing Patterns and Outcomes of Patients With Metastatic Pancreatic Cancer Treated With a Liposomal Irinotecan Regimen in the United States. Pancreas 49, 193–200 (2020).

[35] Hemant Ishwaran, Udaya B. Kogalur, Eugene H. Blackstone & Michael S. Lauer. Random survival forests. The Annals of Applied Statistics 2, 841–860 (2008). URL 10.1214/08-AOAS169.

[36] Katzman, J. L. et al. DeepSurv: personalized treatment recommender system using a Cox proportional hazards deep neural network. BMC Medical Research Methodology 18, 24 (2018). URL 10.1186/s12874-018-0482-1.

[37] Lee, C., Zame, W., Yoon, J. & Schaar, M. v. d. DeepHit: A Deep Learning Approach to Survival Analysis With Competing Risks. Proceedings of the AAAI Conference on Artificial Intelligence 32 (2018). URL https://ojs.aaai.org/index.php/AAAI/article/view/11842.

38. [38] Chen, G. H. An introduction to deep survival analysis models for predicting time-to-event outcomes (2024). URL https://arxiv.org/abs/2410.01086.arXiv:2410.01086.

[39] Antolini, L., Boracchi, P. & Biganzoli, E. A time-dependent discrimination index for survival data. Statistics in Medicine 24, 3927–3944 (2005).

[40] Hung, H. & Chiang, C.-T. Estimation methods for time-dependent auc models with survival data. Canadian Journal of Statistics 38, 8–26 (2010). URL https://onlinelibrary.wiley.com/doi/abs/10.1002/cjs.10046.

[41] Uno, H., Cai, T., Pencina, M. J., D’Agostino, R. B. & Wei, L. J. On the C-statistics for evaluating overall adequacy of risk prediction procedures with censored survival data. Statistics in Medicine 30, 1105–1117 (2011).

[42] Graf, E., Schmoor, C., Sauerbrei, W. & Schumacher, M. Assessment and comparison of prognostic classification schemes for survival data. Statistics in Medicine 18, 2529–2545 (1999). URL https://onlinelibrary.wiley.com/doi/abs/10.1002/%28SICI%291097-0258%2819990915/30%2918%3A17/18%3C2529%3A%3AAID-SIM274%3E3.0.CO%3B2-5.

[43] BioRender.com. Figures created with BioRender. https://BioRender.com/56dcnu2 (2026). Accessed 11 Feb 2026.

