## Supplement for "Reconstructing real-world metastatic lines of therapy enables progression risk stratification in breast cancer"

### Supplementary Information

#### Tables

**Supplementary Table 1**: Supplementary tables for the breast cancer subset.

(a) Baseline characteristics of the breast cancer subset.

| **Variable** | **Overall** |
|---|---|
| n | 2,881 |
| Age at first modeled line initiation, mean (sd) | 55.5 (12.3) |
| *Stage at diagnosis* | |
| Stage I–III | 2,047 (71.1%) |
| Stage IV | 834 (28.9%) |

(b) Patient-level HR/HER2 status by age at the first retained modeled line (row-wise %).

| **Age group** | **HR+** | **HR−** | **HER2+** | **HER2−** |
|---|---|---|---|---|
| ≤40 | 74.4 | 25.6 | 28.6 | 71.4 |
| 41–50 | 79.5 | 20.5 | 21.3 | 78.7 |
| 51–65 | 79.1 | 20.9 | 14.9 | 85.1 |
| 66–75 | 83.6 | 16.4 | 11.5 | 88.5 |
| 76+ | 83.1 | 16.9 | 9.2 | 90.8 |

*Notes:* One record per patient at the first retained modeled line; 2,881 patients in total. Age-group denominators are 360, 625, 1,228, 538, and 130, respectively. Percentages are descriptive; no significance test is reported.

**Supplementary Table 2**: Most common first-line metastatic drug combinations at line initiation (mLoT1), stratified by disease subtypes.

| Subtype | Rank | Regimen | # (%) |
|---|---|---|---|
| HR−/HER2− | 1 | Chemo Capecitabine | 76 (19.5%) |
| | 2 | Chemo Other | 62 (15.9%) |
| | 3 | Chemo Paclitaxel | 53 (13.6%) |
| | 4 | Chemo Other,<br>Chemo Gemcitabine | 33 (8.5%) |
| | 5 | Chemo Other,<br>Chemo Paclitaxel | 23 (5.9%) |
| HR−/HER2+ | 1 | Chemo Paclitaxel,<br>Biologic Pertuzumab,<br>Biologic Trastuzumab | 54 (38.3%) |
| | 2 | Chemo Doxorubicin,<br>Chemo Cyclophosphamide | 8 (5.7%) |
| | 3 | Biologic Trastuzumab | 7 (5.0%) |
| | 4 | Biologic Pertuzumab,<br>Biologic Trastuzumab | 6 (4.3%) |
| | 5 | Biologic Ado-Trastuzumab<br>Emtansine | 6 (4.3%) |
| HR+/HER2− | 1 | Hormone Letrozole,<br>Targeted Palbociclib | 196 (12.5%) |
| | 2 | Hormone Letrozole | 137 (8.8%) |
| | 3 | Hormone Other,<br>Hormone Letrozole,<br>Targeted Palbociclib | 81 (5.2%) |
| | 4 | Chemo Capecitabine | 77 (4.9%) |
| | 5 | Hormone Fulvestrant | 75 (4.8%) |
| HR+/HER2+ | 1 | Chemo Paclitaxel,<br>Biologic Pertuzumab,<br>Biologic Trastuzumab | 41 (16.6%) |
| | 2 | Hormone Letrozole | 13 (5.3%) |
| | 3 | Biologic Ado-Trastuzumab<br>Emtansine | 12 (4.9%) |
| | 4 | Chemo Other,<br>Chemo Paclitaxel,<br>Biologic Pertuzumab,<br>Biologic Trastuzumab | 10 (4.0%) |
| | 5 | Chemo Paclitaxel,<br>Biologic Trastuzumab,<br>Biologic Investigative | 9 (3.6%) |

*Notes:* Percentages reflect the share within each HR/HER2 subtype. Drug names are line-broken within cells for readability.

**Supplementary Table 3**: Most common mLoT2+ treatment-category combinations at mLoT initiation, stratified by disease subtypes.

| Subtype | Rank | Regimen | # (%) |
|---|---|---|---|
| HR−/HER2− | 1 | Chemo | 418 (65.0%) |
| | 2 | Biologic | 70 (10.9%) |
| | 3 | Chemo, Biologic | 40 (6.2%) |
| | 4 | Chemo, Immuno | 34 (5.3%) |
| | 5 | Chemo, Targeted | 29 (4.5%) |
| HR−/HER2+ | 1 | Chemo, Biologic | 99 (33.4%) |
| | 2 | Biologic | 82 (27.7%) |
| | 3 | Chemo, Targeted, Biologic | 45 (15.2%) |
| | 4 | Targeted, Biologic | 20 (6.8%) |
| | 5 | Chemo | 20 (6.8%) |
| HR+/HER2− | 1 | Chemo | 1,940 (42.5%) |
| | 2 | Hormone, Targeted | 1,274 (27.9%) |
| | 3 | Hormone | 545 (11.9%) |
| | 4 | Chemo, Targeted | 262 (5.7%) |
| | 5 | Biologic | 191 (4.2%) |
| HR+/HER2+ | 1 | Chemo, Biologic | 254 (26.9%) |
| | 2 | Hormone, Biologic | 186 (19.7%) |
| | 3 | Biologic | 136 (14.4%) |
| | 4 | Chemo | 64 (6.8%) |
| | 5 | Chemo, Targeted, Biologic | 61 (6.5%) |

*Notes:* Percentages reflect the share within each HR/HER2 subtype among mLoT2+ lines.

**Supplementary Table 4**: Per-horizon performance across models for PFS prediction. Values are mean ± std over splits. Requested horizons [90, 180, 365, 730] resolve to [90, 180, 365, 723] on the common evaluation grid. For Brier score, lower is better.

(a) IPCW-$C$

| **Model** | **90 d** | **180 d** | **365 d** | **730 d** |
|---|---|---|---|---|
| CoxPH | 0.691 ± 0.008 | 0.682 ± 0.007 | 0.681 ± 0.007 | 0.679 ± 0.007 |
| DeepHit | 0.696 ± 0.008 | 0.683 ± 0.007 | 0.675 ± 0.009 | 0.651 ± 0.011 |
| DeepSurv | 0.694 ± 0.010 | 0.683 ± 0.009 | 0.682 ± 0.010 | 0.680 ± 0.009 |
| GBSA | 0.693 ± 0.006 | 0.684 ± 0.006 | 0.683 ± 0.006 | 0.681 ± 0.006 |
| RSF | 0.684 ± 0.006 | 0.674 ± 0.006 | 0.671 ± 0.007 | 0.666 ± 0.007 |

(b) AUC

| **Model** | **90 d** | **180 d** | **365 d** | **730 d** |
|---|---|---|---|---|
| CoxPH | 0.730 ± 0.008 | 0.759 ± 0.011 | 0.818 ± 0.008 | 0.842 ± 0.013 |
| DeepHit | 0.736 ± 0.010 | 0.759 ± 0.012 | 0.811 ± 0.008 | 0.806 ± 0.026 |
| DeepSurv | 0.732 ± 0.012 | 0.760 ± 0.011 | 0.816 ± 0.012 | 0.841 ± 0.015 |
| GBSA | 0.733 ± 0.007 | 0.763 ± 0.011 | 0.824 ± 0.008 | 0.847 ± 0.012 |
| RSF | 0.724 ± 0.008 | 0.753 ± 0.012 | 0.814 ± 0.009 | 0.845 ± 0.012 |

(c) Brier score

| **Model** | **90 d** | **180 d** | **365 d** | **730 d** |
|---|---|---|---|---|
| CoxPH | 0.202 ± 0.005 | 0.188 ± 0.005 | 0.122 ± 0.006 | 0.075 ± 0.007 |
| DeepHit | 0.221 ± 0.004 | 0.210 ± 0.006 | 0.137 ± 0.009 | 0.087 ± 0.011 |
| DeepSurv | 0.201 ± 0.006 | 0.187 ± 0.006 | 0.121 ± 0.007 | 0.075 ± 0.007 |
| GBSA | 0.201 ± 0.004 | 0.186 ± 0.005 | 0.119 ± 0.006 | 0.074 ± 0.007 |
| RSF | 0.204 ± 0.005 | 0.189 ± 0.006 | 0.121 ± 0.006 | 0.074 ± 0.007 |

**Supplementary Table 5**: Feature-level baseline value contrasts between high- and low-risk tertiles (GBSA). Cohen's *d* (H–L) is computed on cohort-centered baseline feature values within each HR/HER2 subtype and line group. Displayed rows show the top five cohort-specific contrasts after setting aside the recurrent cross-subgroup features *first metastatic line* and *genomic data missing*; when present, those recurring effects are reported in the note below.

| **Cohort** | **Top 5 baseline features (Cohen's $d$, H–L)** |
|---|---|
| HR−/HER2− | TP53 mutation (+1.240)<br>Chemotherapy: non-top-5 agents (+1.182)<br>Liver mets in last 90d (+1.180)<br>Liver mets ever (+1.126)<br>Cancer reported; screened chest (+1.012) |
| HR−/HER2+ | Cancer reported; screened abdomen (+1.524)<br>Cancer reported; screened pelvis (+1.524)<br>Cancer reported; screened chest (+1.490)<br>TP53 mutation (+1.400)<br>Recent imaging, chest: cancer present (+1.080) |
| HR+/HER2− | Liver mets in last 90d (+1.788)<br>Liver mets ever (+1.780)<br>CA15–3 > 30 U/mL (last value) (+1.369)<br>Chemotherapy: non-top-5 agents (+1.307)<br>CA15–3 max (last 180d) (+1.235) |
| HR+/HER2+ | Liver mets in last 90d (+1.429)<br>CA15–3 > 30 U/mL (last value) (+1.224)<br>Cancer reported; screened chest (+1.101)<br>Cancer reported; screened pelvis (+1.081)<br>CEA > 5 ng/mL (last value) (+1.077) |
| mLoT1 | No CEA in last 180d (−1.088)<br>No CA15–3 in last 180d (−1.078)<br>TP53 mutation (+1.022)<br>No CEA in last 60d (−1.020)<br>No CA15–3 in last 60d (−1.009) |
| mLoT2+ | Liver mets in last 90d (+1.671)<br>Chemotherapy: non-top-5 agents (+1.457)<br>Liver mets ever (+1.192)<br>CA15–3 max (last 180d) (+1.117)<br>CA15–3 last value (log) (+1.076) |

*Recurring excluded contrasts:* HR−/HER2− first metastatic line (−1.689), genomic data missing (−1.411); HR−/HER2+ genomic data missing (−1.685), first metastatic line (−1.507); HR+/HER2− first metastatic line (−1.705), genomic data missing (−1.644); HR+/HER2+ first metastatic line (−1.276), genomic data missing (−0.942); mLoT1 genomic data missing (−0.983); mLoT2+ genomic data missing (−0.873).

**Supplementary Table 6**: Group-level SHAP contribution contrasts between high- and low-risk tertiles (GBSA). Cohen's $d$ (H–L) is computed on group share of total absolute SHAP attribution within each cohort.

| **Cohort** | **Top 5 feature groups** | **Cohen's $d$ (H–L)** |
|---|---|---|
| HR−/HER2− | Tumor marker kinetics | −1.156 |
| | Imaging coverage and cancer status | −0.986 |
| | Genomics and therapeutic targets | +0.910 |
| | Measurement cadence | −0.805 |
| | Treatment exposure | +0.541 |
| HR−/HER2+ | Genomics and therapeutic targets | +1.184 |
| | Imaging coverage and cancer status | −0.846 |
| | Recent ECOG status | +0.728 |
| | Measurement cadence | −0.427 |
| | Tumor marker kinetics | −0.409 |
| HR+/HER2− | Measurement cadence | −0.841 |
| | Imaging coverage and cancer status | −0.833 |
| | Genomics and therapeutic targets | +0.669 |
| | Recent ECOG status | +0.560 |
| | Initial diagnosis | −0.408 |
| HR+/HER2+ | Imaging coverage and cancer status | −0.640 |
| | Recent ECOG status | +0.558 |
| | Tumor site burden | +0.445 |
| | Tumor marker kinetics | −0.311 |
| | Treatment exposure | −0.203 |
| mLoT1 | Genomics and therapeutic targets | +1.114 |
| | Recent ECOG status | +0.853 |
| | Treatment exposure | −0.653 |
| | Tumor marker kinetics | −0.511 |
| | Initial diagnosis | +0.349 |
| mLoT2+ | Imaging coverage and cancer status | −0.816 |
| | Initial diagnosis | −0.799 |
| | Treatment exposure | +0.747 |
| | Measurement cadence | −0.665 |
| | Tumor marker kinetics | −0.415 |

**Supplementary Table 7**: Descriptive mean performance differences across five outer test folds, without versus with treatment masking. Δ is calculated as (no masking) − (masking). These fold means are not pooled out-of-fold differences; no significance test across folds is reported.

| model | $\Delta C$ | Δ IBS | Δ AUC |
|---|---|---|---|
| coxph | 0.012541 | -0.005267 | 0.016274 |
| deephit | 0.012488 | -0.001671 | 0.016130 |
| deepsurv | 0.014982 | -0.005888 | 0.019954 |
| gbsa | 0.010389 | -0.004447 | 0.013161 |
| rsf | 0.004182 | -0.002841 | 0.006082 |

**Supplementary Table 8**: Global performance metrics without late-start treatment masking.

| **Metric** | **CoxPH** | **DeepHit** | **DeepSurv** | **GBSA** | **RSF** |
|---|---|---|---|---|---|
| C-index | 0.691 ± 0.007 | **0.697 ± 0.008** | 0.695 ± 0.006 | 0.692 ± 0.007 | 0.678 ± 0.005 |
| mean AUC | 0.770 ± 0.008 | **0.775 ± 0.008** | 0.775 ± 0.006 | 0.770 ± 0.007 | 0.753 ± 0.005 |
| AUC @ 1y | 0.840 ± 0.010 | 0.831 ± 0.009 | 0.838 ± 0.005 | **0.842 ± 0.010** | 0.826 ± 0.006 |
| AUC @ 2y | 0.860 ± 0.012 | 0.827 ± 0.026 | 0.856 ± 0.014 | **0.864 ± 0.012** | 0.859 ± 0.010 |
| IBS | 0.120 ± 0.004 | 0.139 ± 0.007 | **0.120 ± 0.004** | 0.120 ± 0.004 | 0.123 ± 0.004 |

**Supplementary Table 9**: Descriptive mean performance differences across five outer test folds, with age excluded versus included. Δ is calculated as (no age) − (age). These fold means are not pooled out-of-fold differences; no significance test across folds is reported.

| model | $\Delta C$ | Δ IBS | Δ AUC |
|---|---|---|---|
| coxph | -0.001281 | 0.000178 | -0.001744 |
| deephit | -0.008931 | 0.005091 | -0.011663 |
| deepsurv | -0.001944 | -0.000017 | -0.002720 |
| gbsa | -0.001095 | 0.000001 | -0.001501 |
| rsf | 0.000242 | -0.000417 | 0.000824 |

**Supplementary Table 10**: Global performance metrics with age excluded from baseline covariates (age ablation).

| Metric | CoxPH | DeepHit | DeepSurv | GBSA | RSF |
|---|---|---|---|---|---|
| C-index | $0.678 \pm 0.007$ | $0.676 \pm 0.011$ | $0.678 \pm 0.009$ | $\mathbf{0.680 \pm 0.006}$ | $0.675 \pm 0.007$ |
| mean AUC | $0.752 \pm 0.008$ | $0.747 \pm 0.015$ | $0.752 \pm 0.012$ | $\mathbf{0.755 \pm 0.007}$ | $0.748 \pm 0.006$ |
| AUC @ 1y | $0.818 \pm 0.008$ | $0.802 \pm 0.015$ | $0.815 \pm 0.011$ | $\mathbf{0.824 \pm 0.007}$ | $0.814 \pm 0.008$ |
| AUC @ 2y | $0.842 \pm 0.013$ | $0.783 \pm 0.043$ | $0.840 \pm 0.015$ | $\mathbf{0.848 \pm 0.014}$ | $0.847 \pm 0.013$ |
| IBS | $0.126 \pm 0.004$ | $0.145 \pm 0.009$ | $0.125 \pm 0.005$ | $\mathbf{0.124 \pm 0.004}$ | $0.125 \pm 0.004$ |

**Supplementary Table 11**: Global performance metrics under the 14-day treatment-window sensitivity analysis.

| Metric | CoxPH | DeepHit | DeepSurv | GBSA | RSF |
|---|---|---|---|---|---|
| C-index | $0.677 \pm 0.006$ | $\mathbf{0.682 \pm 0.006}$ | $0.679 \pm 0.006$ | $0.680 \pm 0.006$ | $0.672 \pm 0.006$ |
| mean AUC | $0.750 \pm 0.007$ | $\mathbf{0.754 \pm 0.007}$ | $0.753 \pm 0.007$ | $0.753 \pm 0.008$ | $0.742 \pm 0.009$ |
| AUC @ 1y | $0.814 \pm 0.008$ | $0.811 \pm 0.008$ | $0.816 \pm 0.012$ | $\mathbf{0.819 \pm 0.009}$ | $0.807 \pm 0.007$ |
| AUC @ 2y | $0.839 \pm 0.029$ | $0.812 \pm 0.030$ | $0.843 \pm 0.024$ | $\mathbf{0.845 \pm 0.027}$ | $0.838 \pm 0.025$ |
| IBS | $0.128 \pm 0.002$ | $0.144 \pm 0.002$ | $0.127 \pm 0.003$ | $\mathbf{0.127 \pm 0.002}$ | $0.129 \pm 0.001$ |

**Supplementary Table 12**: Global performance metrics under the 42-day treatment-window sensitivity analysis.

| Metric | CoxPH | DeepHit | DeepSurv | GBSA | RSF |
|---|---|---|---|---|---|
| C-index | $0.677 \pm 0.008$ | $\mathbf{0.680 \pm 0.007}$ | $0.680 \pm 0.007$ | $0.680 \pm 0.007$ | $0.674 \pm 0.006$ |
| mean AUC | $0.755 \pm 0.009$ | $0.757 \pm 0.008$ | $0.758 \pm 0.010$ | $\mathbf{0.758 \pm 0.008}$ | $0.750 \pm 0.008$ |
| AUC @ 1y | $0.815 \pm 0.006$ | $0.811 \pm 0.007$ | $0.817 \pm 0.006$ | $\mathbf{0.821 \pm 0.007}$ | $0.812 \pm 0.009$ |
| AUC @ 2y | $0.841 \pm 0.013$ | $0.812 \pm 0.017$ | $0.843 \pm 0.014$ | $\mathbf{0.848 \pm 0.012}$ | $0.844 \pm 0.011$ |
| IBS | $0.126 \pm 0.004$ | $0.141 \pm 0.006$ | $\mathbf{0.124 \pm 0.004}$ | $0.124 \pm 0.004$ | $0.125 \pm 0.004$ |

**Supplementary Table 13**: Global performance metrics from pooled out-of-fold predictions for all 8,791 modeled mLoTs from 2,881 patients, with 95% percentile confidence intervals from 1,000 patient-level bootstrap resamples (each sampled patient contributes all of their mLoTs; predictions are held fixed, so intervals reflect uncertainty in the evaluation sample, not in model training). For DeepHit, pooled predictions are first reconstructed on a common daily grid using the same piecewise-linear survival interpolation used in fold-level evaluation.

| Metric | CoxPH | DeepHit | DeepSurv | GBSA | RSF |
|---|---|---|---|---|---|
| C-index | 0.678 (0.672, 0.685) | **0.683 (0.676, 0.690)** | 0.678 (0.671, 0.685) | 0.681 (0.674, 0.687) | 0.673 (0.667, 0.680) |
| mean AUC | 0.754 (0.745, 0.762) | **0.757 (0.748, 0.766)** | 0.753 (0.744, 0.762) | **0.757 (0.748, 0.765)** | 0.746 (0.737, 0.755) |
| AUC @ 1y | 0.818 (0.806, 0.829) | 0.811 (0.800, 0.822) | 0.814 (0.802, 0.826) | **0.824 (0.812, 0.835)** | 0.813 (0.802, 0.824) |
| AUC @ 2y | 0.843 (0.828, 0.856) | 0.799 (0.782, 0.817) | 0.842 (0.826, 0.855) | **0.848 (0.834, 0.861)** | 0.845 (0.830, 0.859) |
| IBS | 0.126 (0.122, 0.129) | 0.140 (0.136, 0.145) | 0.125 (0.122, 0.129) | **0.124 (0.121, 0.128)** | 0.126 (0.123, 0.129) |

**Supplementary Table 14**: Best-performing model configurations selected on outer folds, with selection counts. These configurations are reused in the ablation experiments.

| Model | Selected configurations (count) |
|---|---|
| CoxPH | `alpha-727.895` (4)<br>`alpha-529.832` (1) |
| DeepHit | `dh104-base-strongreg` (5) |
| DeepSurv | `ds-lite-64-strongreg` (5) |
| GBSA | `gbsa-simple-400` (5) |
| RSF | `rsf-mid-400` (3)<br>`rsf-simple-400` (2) |

**Supplementary Table 15**: Sensitivity to the QC exclusions. MSK-CHORD saved GBSA models evaluated on all 11,085 eligible lines, including the 2,294 lines removed by QC, with pre-QC PFS labels restored. The models were trained on the original retained cohort and were not refitted. Retained lines keep their original out-of-fold predictions. For the 1,076 patients represented in the retained cohort but who also have QC lines, all restored lines were evaluated with the saved outer-fold model. Each of the 293 patients absent from the retained cohort was assigned to a single outer fold with a fixed seed, independently of outcome, and all lines of that patient are scored. C is pooled adjusted-Antolini concordance; IBS is descriptive and uses the original cohort's censoring times from day 28 to day 730, with censoring weights estimated within each evaluated cohort. Values are point estimates.

| Cohort | Lines | Patients | C | IBS |
|---|---|---|---|---|
| Original retained cohort | 8,791 | 2,881 | 0.6807 | 0.1241 |
| All eligible lines, including QC exclusions | 11,085 | 3,174 | 0.6815 | 0.1258 |
| QC-excluded lines only | 2,294 | 1,369 | 0.6953 | 0.1324 |

**Supplementary Table 16**: Feature-group selection rules used for grouped attribution analyses. Each group maps baseline covariates at line initiation into a clinically interpretable domain.

<table>
<tr><th>Feature group</th><th>Selection rule</th></tr>
<tr><td>Tumor marker kinetics</td><td>Prefixes CA15_3_ and CEA_; exclude regex _MISSING$ and _LAST_OBS_DAY$.</td></tr>
<tr><td>Recent ECOG status</td><td>Exact features ECOG_LAST_OBS and EVER_GE2_180D.</td></tr>
<tr><td>Tumor site burden</td><td>Regex ^TUMOR_SITE_.*_(IN_WINDOW|EVER)$.</td></tr>
<tr><td>Genomics and therapeutic targets</td><td>Exact features HR and HER2; prefixes PDL1, MMR, and GENOMICS; exclude exact feature GENOMICS_MISSING.</td></tr>
<tr><td>Initial diagnosis</td><td>Prefixes STAGE, CLINICAL_GROUP, PATH_GROUP, HISTOLOGIC, SUMMARY, CANCER_SITE_SUBSITE, AGE, and GENDER_IS_FEMALE.</td></tr>
<tr><td>Treatment exposure</td><td>Prefixes PLANNED_ and IS_MLOT.</td></tr>
<tr><td>Imaging coverage and cancer status</td><td>Regex ^CANCER_.*_IMAGED_(Y|N|INDET)_STATUS$ and ^CANCER_.*_EVER$; exclude regex ^CANCER_SITE_SUBSITE_.</td></tr>
<tr><td>Measurement cadence</td><td>Exact features ECOG_LAST_OBS_DAY, ECOG_MISSING, and GENOMICS_MISSING; regex ^CANCER_.*_MISSING$, ^CA15_3_.*_MISSING$, ^CEA_.*_MISSING$, ^CA15_3_LAST_OBS_DAY$, and ^CEA_LAST_OBS_DAY$.</td></tr>
</table>

**Supplementary Table 17**: External evaluation on AACR Project GENIE BPC metastatic breast-cancer regimens at DFCI and VICC. The upper block uses released imaging-based PFS (PFS-I: Progressing or death); the lower block (PFS-I + Mixed, used in the main text) also counts Mixed imaging assessments as progression. Each endpoint applies the 28-day minimum duration and 730-day administrative censoring. AUC and IBS use inverse-probability-of-censoring weights estimated from each site's own regimens. Metrics are mean ± SD across the same five fixed MSK-CHORD outer-fold models. SD describes model variation and is not a patient-level confidence interval.

| Institution | Regimens | Patients | Events | C | AUC | AUC@1y | AUC@2y | IBS |
|---|---|---|---|---|---|---|---|---|
| **Released PFS-I: Progressing or death** | | | | | | | | |
| DFCI | 1,398 | 317 | 942 | $0.643 \pm 0.002$ | $0.707 \pm 0.003$ | $0.731 \pm 0.006$ | $0.755 \pm 0.010$ | $0.171 \pm 0.001$ |
| VICC | 701 | 153 | 460 | $0.626 \pm 0.006$ | $0.691 \pm 0.007$ | $0.750 \pm 0.008$ | $0.780 \pm 0.009$ | $0.163 \pm 0.001$ |
| **PFS-I + Mixed (main analysis): Progressing or Mixed, or death** | | | | | | | | |
| DFCI | 1,394 | 317 | 987 | $0.649 \pm 0.002$ | $0.714 \pm 0.003$ | $0.737 \pm 0.006$ | $0.770 \pm 0.010$ | $0.166 \pm 0.001$ |
| VICC | 695 | 152 | 485 | $0.634 \pm 0.006$ | $0.700 \pm 0.007$ | $0.758 \pm 0.009$ | $0.799 \pm 0.010$ | $0.159 \pm 0.001$ |

*Endpoint and cohort note:* Both blocks count curated metastatic regimens. The 28-day minimum duration and 730-day administrative censoring are applied separately to each endpoint. The PFS-I + Mixed endpoint excludes four additional DFCI and six additional VICC regimens with PFS below 28 days. Both blocks use the same five fixed MSK-CHORD models; values are mean ± SD across models, not ensemble estimates or patient-level confidence intervals.

**Supplementary Table 18**: Complete model input list.

| Input name | Feature group | SP |
|---|---|---|
| **Treatment (36 inputs)** | | |
| PLANNED_AGENT_HORMONE_FULVESTRANT | TREATMENT | No |
| PLANNED_AGENT_CHEMO_OTHER | TREATMENT | No |
| PLANNED_AGENT_CHEMO_CAPECITABINE | TREATMENT | No |
| PLANNED_AGENT_HORMONE_EXEMESTANE | TREATMENT | No |
| PLANNED_AGENT_TARGETED_INVESTIGATIVE | TREATMENT | No |
| PLANNED_AGENT_BONE TREATMENT_ZOLEDRONIC ACID | TREATMENT | No |
| PLANNED_AGENT_CHEMO_PACLITAXEL | TREATMENT | No |
| PLANNED_AGENT_CHEMO_GEMCITABINE | TREATMENT | No |
| PLANNED_AGENT_HORMONE_OTHER | TREATMENT | No |
| PLANNED_AGENT_HORMONE_LETROZOLE | TREATMENT | No |
| PLANNED_AGENT_BIOLOGIC_ADO-TRASTUZUMAB EMTANSINE | TREATMENT | No |
| PLANNED_AGENT_CHEMO_DOXORUBICIN | TREATMENT | No |
| PLANNED_AGENT_BIOLOGIC_PERTUZUMAB | TREATMENT | No |
| PLANNED_AGENT_BIOLOGIC_TRASTUZUMAB | TREATMENT | No |
| PLANNED_AGENT_TARGETED_OTHER | TREATMENT | No |
| PLANNED_AGENT_OTHER_INVESTIGATIVE | TREATMENT | No |
| PLANNED_AGENT_TARGETED_ABEMACICLIB | TREATMENT | No |
| PLANNED_AGENT_TARGETED_EVEROLIMUS | TREATMENT | No |
| PLANNED_AGENT_HORMONE_TAMOXIFEN | TREATMENT | No |
| PLANNED_AGENT_TARGETED_PALBOCICLIB | TREATMENT | No |
| PLANNED_AGENT_BIOLOGIC_INVESTIGATIVE | TREATMENT | No |
| PLANNED_AGENT_CHEMO_CYCLOPHOSPHAMIDE | TREATMENT | No |
| PLANNED_AGENT_HORMONE_ANASTROZOLE | TREATMENT | No |
| PLANNED_AGENT_IMMUNO_PEMBROLIZUMAB | TREATMENT | No |
| PLANNED_AGENT_BIOLOGIC_OTHER | TREATMENT | No |
| PLANNED_AGENT_IMMUNO_INVESTIGATIVE | TREATMENT | No |
| PLANNED_AGENT_TARGETED_ALPELISIB | TREATMENT | No |
| PLANNED_AGENT_BIOLOGIC_FAM-TRASTUZUMAB DERUXTECAN | TREATMENT | No |
| PLANNED_AGENT_BONE TREATMENT_PAMIDRONATE | TREATMENT | No |
| PLANNED_AGENT_OTHER_LETROZOLE-RIBOCICLIB | TREATMENT | No |
| PLANNED_AGENT_IMMUNO_ATEZOLIZUMAB | TREATMENT | No |
| PLANNED_AGENT_IMMUNO_IPILIMUMAB | TREATMENT | No |
| PLANNED_AGENT_IMMUNO_NIVOLUMAB | TREATMENT | No |
| PLANNED_SURGERY | LOCAL_TREATMENT | No |
| PLANNED_RADIATION_THERAPY | LOCAL_TREATMENT | No |
| IS_MLOT1 | TREATMENT | No |
| **Genomics (52 inputs)** | | |
| PDL1_POS | PDL1 | No |
| PDL1_NEG | PDL1 | No |
| PDL1_UNKNOWN | PDL1 | No |
| MMR_NEG | MMR | No |
| MMR_POS | MMR | No |
| MMR_UNKNOWN | MMR | No |
| GENOMICS_MUTATION_AKT1 | GENOMICS | No |
| GENOMICS_MUTATION_ARID1A | GENOMICS | No |
| GENOMICS_MUTATION_ATM | GENOMICS | No |
| GENOMICS_MUTATION_BRCA1 | GENOMICS | No |
| GENOMICS_MUTATION_BRCA2 | GENOMICS | No |
| GENOMICS_MUTATION_CBFB | GENOMICS | No |

Continued on next page.

**Supplementary Table 18**: Complete model input list (continued).

| Input name | Feature group | SP |
|---|---|---|
| GENOMICS_MUTATION_CDH1 | GENOMICS | No |
| GENOMICS_MUTATION_CDKN1B | GENOMICS | No |
| GENOMICS_MUTATION_ESR1 | GENOMICS | No |
| GENOMICS_MUTATION_FOXA1 | GENOMICS | No |
| GENOMICS_MUTATION_GATA3 | GENOMICS | No |
| GENOMICS_MUTATION_KMT2C | GENOMICS | No |
| GENOMICS_MUTATION_KMT2D | GENOMICS | No |
| GENOMICS_MUTATION_MAP2K4 | GENOMICS | No |
| GENOMICS_MUTATION_MAP3K1 | GENOMICS | No |
| GENOMICS_MUTATION_NCOR1 | GENOMICS | No |
| GENOMICS_MUTATION_NF1 | GENOMICS | No |
| GENOMICS_MUTATION_PIK3CA | GENOMICS | No |
| GENOMICS_MUTATION_PIK3R1 | GENOMICS | No |
| GENOMICS_MUTATION_PTEN | GENOMICS | No |
| GENOMICS_MUTATION_RB1 | GENOMICS | No |
| GENOMICS_MUTATION_RUNX1 | GENOMICS | No |
| GENOMICS_MUTATION_SF3B1 | GENOMICS | No |
| GENOMICS_MUTATION_TBX3 | GENOMICS | No |
| GENOMICS_MUTATION_TP53 | GENOMICS | No |
| GENOMICS_AMPLIFICATION_CCND1 | GENOMICS | No |
| GENOMICS_AMPLIFICATION_CCNE1 | GENOMICS | No |
| GENOMICS_AMPLIFICATION_EGFR | GENOMICS | No |
| GENOMICS_AMPLIFICATION_ERBB2 | GENOMICS | No |
| GENOMICS_AMPLIFICATION_FGFR1 | GENOMICS | No |
| GENOMICS_AMPLIFICATION_FGFR2 | GENOMICS | No |
| GENOMICS_AMPLIFICATION_FOXA1 | GENOMICS | No |
| GENOMICS_AMPLIFICATION_IGF1R | GENOMICS | No |
| GENOMICS_AMPLIFICATION_MCL1 | GENOMICS | No |
| GENOMICS_AMPLIFICATION_MYC | GENOMICS | No |
| GENOMICS_AMPLIFICATION_PAK1 | GENOMICS | No |
| GENOMICS_AMPLIFICATION_PIK3CA | GENOMICS | No |
| GENOMICS_DELETION_ATM | GENOMICS | No |
| GENOMICS_DELETION_BRCA1 | GENOMICS | No |
| GENOMICS_DELETION_BRCA2 | GENOMICS | No |
| GENOMICS_DELETION_CDKN2A | GENOMICS | No |
| GENOMICS_DELETION_CDKN2B | GENOMICS | No |
| GENOMICS_DELETION_MAP2K4 | GENOMICS | No |
| GENOMICS_DELETION_PTEN | GENOMICS | No |
| GENOMICS_DELETION_RB1 | GENOMICS | No |
| GENOMICS_MISSING | GENOMICS | Yes |
| **Clinicopathologic (36 inputs)** | | |
| ECOG_LAST_OBS | ECOG | No |
| ECOG_LAST_OBS_DAY | ECOG | Yes |
| EVER_GE2_180D | ECOG | No |
| ECOG_MISSING | ECOG | Yes |
| GENDER_IS_FEMALE | CLINICAL | No |
| AGE | CLINICAL | No |
| HR | CLINICAL | No |
| HER2 | CLINICAL | No |
| STAGE_CDM_DERIVED_IV | DIAGNOSIS | No |

Continued on next page.

**Supplementary Table 18**: Complete model input list (continued).

| Input name | Feature group | SP |
|---|---|---|
| CLINICAL_GROUP_0 | DIAGNOSIS | No |
| CLINICAL_GROUP_1 | DIAGNOSIS | No |
| CLINICAL_GROUP_2 | DIAGNOSIS | No |
| CLINICAL_GROUP_3 | DIAGNOSIS | No |
| CLINICAL_GROUP_4 | DIAGNOSIS | No |
| CLINICAL_GROUP_8 | DIAGNOSIS | No |
| CLINICAL_GROUP_9 | DIAGNOSIS | No |
| PATH_GROUP_0 | DIAGNOSIS | No |
| PATH_GROUP_1 | DIAGNOSIS | No |
| PATH_GROUP_2 | DIAGNOSIS | No |
| PATH_GROUP_3 | DIAGNOSIS | No |
| PATH_GROUP_4 | DIAGNOSIS | No |
| PATH_GROUP_8 | DIAGNOSIS | No |
| PATH_GROUP_9 | DIAGNOSIS | No |
| HISTOLOGIC_INFILTRATING DUCT CARCINOMA | DIAGNOSIS | No |
| HISTOLOGIC_LOBULAR CA + IFDC/DCIS | DIAGNOSIS | No |
| HISTOLOGIC_LOBULAR CARCINOMA, NOS | DIAGNOSIS | No |
| HISTOLOGIC_OTHER | DIAGNOSIS | No |
| SUMMARY_DISTANT | DIAGNOSIS | No |
| SUMMARY_IN_SITU | DIAGNOSIS | No |
| SUMMARY_LOCALIZED | DIAGNOSIS | No |
| SUMMARY_REGIONAL | DIAGNOSIS | No |
| SUMMARY_UNKNOWN | DIAGNOSIS | No |
| CANCER_SITE_SUBSITE_NOS | DIAGNOSIS | No |
| CANCER_SITE_SUBSITE_OTHER | DIAGNOSIS | No |
| CANCER_SITE_SUBSITE_OVERLAPPING LESION OF BREAST | DIAGNOSIS | No |
| CANCER_SITE_SUBSITE_UOQ | DIAGNOSIS | No |
| **Tumor markers (36 inputs)** | | |
| CA15_3_SHORT_RW_MEAN | TUMOR_MARKERS | No |
| CA15_3_SHORT_MAX | TUMOR_MARKERS | No |
| CA15_3_SHORT_MIN | TUMOR_MARKERS | No |
| CA15_3_SHORT_MISSING | TUMOR_MARKERS | Yes |
| CA15_3_SHORT_SLOPE | TUMOR_MARKERS | No |
| CA15_3_SHORT_DELTA_LAST | TUMOR_MARKERS | No |
| CA15_3_LONG_RW_MEAN | TUMOR_MARKERS | No |
| CA15_3_LONG_MAX | TUMOR_MARKERS | No |
| CA15_3_LONG_MIN | TUMOR_MARKERS | No |
| CA15_3_LONG_MISSING | TUMOR_MARKERS | Yes |
| CA15_3_LONG_SLOPE | TUMOR_MARKERS | No |
| CA15_3_LONG_DELTA_LAST | TUMOR_MARKERS | No |
| CA15_3_LAST_OBS_LOG | TUMOR_MARKERS | No |
| CA15_3_LAST_OBS_DAY | TUMOR_MARKERS | Yes |
| CA15_3_SHORT_OVER_LONG | TUMOR_MARKERS | No |
| CA15_3_SHORT_MINUS_LONG | TUMOR_MARKERS | No |
| CA15_3_ABOVE_CUTOFF | TUMOR_MARKERS | No |
| CA15_3_RISE_GT20PCT | TUMOR_MARKERS | No |
| CEA_SHORT_RW_MEAN | TUMOR_MARKERS | No |
| CEA_SHORT_MAX | TUMOR_MARKERS | No |
| CEA_SHORT_MIN | TUMOR_MARKERS | No |
| CEA_SHORT_MISSING | TUMOR_MARKERS | Yes |

Continued on next page.

**Supplementary Table 18**: Complete model input list (continued).

| Input name | Feature group | SP |
|---|---|---|
| CEA_SHORT_SLOPE | TUMOR_MARKERS | No |
| CEA_SHORT_DELTA_LAST | TUMOR_MARKERS | No |
| CEA_LONG_RW_MEAN | TUMOR_MARKERS | No |
| CEA_LONG_MAX | TUMOR_MARKERS | No |
| CEA_LONG_MIN | TUMOR_MARKERS | No |
| CEA_LONG_MISSING | TUMOR_MARKERS | Yes |
| CEA_LONG_SLOPE | TUMOR_MARKERS | No |
| CEA_LONG_DELTA_LAST | TUMOR_MARKERS | No |
| CEA_LAST_OBS_LOG | TUMOR_MARKERS | No |
| CEA_LAST_OBS_DAY | TUMOR_MARKERS | Yes |
| CEA_SHORT_OVER_LONG | TUMOR_MARKERS | No |
| CEA_SHORT_MINUS_LONG | TUMOR_MARKERS | No |
| CEA_ABOVE_CUTOFF | TUMOR_MARKERS | No |
| CEA_RISE_GT20PCT | TUMOR_MARKERS | No |
| **Radiology (45 inputs)** | | |
| CANCER_CHEST_IMAGED_Y_STATUS | CANCER_PRESENCE | Yes |
| CANCER_CHEST_IMAGED_N_STATUS | CANCER_PRESENCE | Yes |
| CANCER_CHEST_IMAGED_INDET_STATUS | CANCER_PRESENCE | Yes |
| CANCER_CHEST_MISSING | CANCER_PRESENCE | Yes |
| CANCER_CHEST_EVER | CANCER_PRESENCE | No |
| CANCER_ABDOMEN_IMAGED_Y_STATUS | CANCER_PRESENCE | Yes |
| CANCER_ABDOMEN_IMAGED_N_STATUS | CANCER_PRESENCE | Yes |
| CANCER_ABDOMEN_IMAGED_INDET_STATUS | CANCER_PRESENCE | Yes |
| CANCER_ABDOMEN_MISSING | CANCER_PRESENCE | Yes |
| CANCER_ABDOMEN_EVER | CANCER_PRESENCE | No |
| CANCER_PELVIS_IMAGED_Y_STATUS | CANCER_PRESENCE | Yes |
| CANCER_PELVIS_IMAGED_N_STATUS | CANCER_PRESENCE | Yes |
| CANCER_PELVIS_IMAGED_INDET_STATUS | CANCER_PRESENCE | Yes |
| CANCER_PELVIS_MISSING | CANCER_PRESENCE | Yes |
| CANCER_PELVIS_EVER | CANCER_PRESENCE | No |
| CANCER_HEAD_IMAGED_Y_STATUS | CANCER_PRESENCE | Yes |
| CANCER_HEAD_IMAGED_N_STATUS | CANCER_PRESENCE | Yes |
| CANCER_HEAD_IMAGED_INDET_STATUS | CANCER_PRESENCE | Yes |
| CANCER_HEAD_MISSING | CANCER_PRESENCE | Yes |
| CANCER_HEAD_EVER | CANCER_PRESENCE | No |
| CANCER_OTHER_IMAGED_Y_STATUS | CANCER_PRESENCE | Yes |
| CANCER_OTHER_IMAGED_N_STATUS | CANCER_PRESENCE | Yes |
| CANCER_OTHER_IMAGED_INDET_STATUS | CANCER_PRESENCE | Yes |
| CANCER_OTHER_MISSING | CANCER_PRESENCE | Yes |
| CANCER_OTHER_EVER | CANCER_PRESENCE | No |
| TUMOR_SITE_ADRENAL GLANDS_IN_WINDOW | TUMOR_SITES | No |
| TUMOR_SITE_ADRENAL GLANDS_EVER | TUMOR_SITES | No |
| TUMOR_SITE_BONE_IN_WINDOW | TUMOR_SITES | No |
| TUMOR_SITE_BONE_EVER | TUMOR_SITES | No |
| TUMOR_SITE_CNS/BRAIN_IN_WINDOW | TUMOR_SITES | No |
| TUMOR_SITE_CNS/BRAIN_EVER | TUMOR_SITES | No |
| TUMOR_SITE_INTRA-ABDOMINAL_IN_WINDOW | TUMOR_SITES | No |
| TUMOR_SITE_INTRA-ABDOMINAL_EVER | TUMOR_SITES | No |
| TUMOR_SITE_LIVER_IN_WINDOW | TUMOR_SITES | No |
| TUMOR_SITE_LIVER_EVER | TUMOR_SITES | No |

Continued on next page.

**Supplementary Table 18**: Complete model input list (continued).

| Input name | Feature group | SP |
|---|---|---|
| TUMOR_SITE_LUNG_IN_WINDOW | TUMOR_SITES | No |
| TUMOR_SITE_LUNG_EVER | TUMOR_SITES | No |
| TUMOR_SITE_LYMPH NODES_IN_WINDOW | TUMOR_SITES | No |
| TUMOR_SITE_LYMPH NODES_EVER | TUMOR_SITES | No |
| TUMOR_SITE_OTHER_IN_WINDOW | TUMOR_SITES | No |
| TUMOR_SITE_OTHER_EVER | TUMOR_SITES | No |
| TUMOR_SITE_PLEURA_IN_WINDOW | TUMOR_SITES | No |
| TUMOR_SITE_PLEURA_EVER | TUMOR_SITES | No |
| TUMOR_SITE_REPRODUCTIVE ORGANS_IN_WINDOW | TUMOR_SITES | No |
| TUMOR_SITE_REPRODUCTIVE ORGANS_EVER | TUMOR_SITES | No |

*Notes:* There are 205 model inputs: 36 treatment, 52 genomic, 36 clinicopathologic, 36 tumor-marker, and 45 radiology inputs. SP indicates membership in the surveillance-pattern ablation. Its 29 inputs overlap the five modalities and do not increase the total. The genomic inputs include 25 mutation, 12 amplification, and eight deletion indicators, six PD-L1/MMR indicators, and one genomic-missingness indicator. Names match the retained model-input columns; candidate genes absent from the final matrix are not additional inputs.

**Supplementary Table 19**: Agreement of the mLoT reconstruction with curated GENIE BPC regimens at DFCI, VICC and MSK, using BPC PFS-I + Mixed (Progressing or Mixed imaging, or death). A reconstructed episode and a BPC regimen match when they belong to the same patient, have the same named non-GnRH drug set and start within 14 days (one-to-one). Regimen agreement uses all retained episodes; endpoint agreement uses matched pairs; both PFS times are capped at 730 days. **a**, Patient and episode flow; patients, formed episodes, retained episodes and labelled BPC regimens are distinct units. **b**, Agreement with conditional denominators; Cohen's kappa is descriptive. **c**, Mutually exclusive regimen disagreement categories. **d**, Endpoint disagreement categories among same-regimen pairs, assigned to the first applicable rule difference; rows 3 and 4 together are the censoring-rule differences; these categories are not a corrected validation score.

**a. Patient and episode flow**

| Step | DFCI | VICC | MSK |
|---|---|---|---|
| GENIE BPC breast cancer patients | 428 | 173 | 529 |
| Index cancer is breast carcinoma (breast sarcoma excluded) | 424 | 171 | 509 |
| Metastatic disease recorded (stage IV, or a distant-metastasis date) | 351 | 165 | 361 |
| At least one metastatic-setting drug record and one imaging assessment | 305 | 148 | 326 |
| Treatment episodes formed by our algorithm | 1,012 | 504 | 1,438 |
| Excluded: no imaging report in the 90 days before the episode start | 83 | 34 | 140 |
| Excluded: no imaging assessment or death after the episode start | 36 | 27 | 54 |
| Excluded: PFS shorter than 28 days | 13 | 8 | 28 |
| Excluded by QC: chemotherapy and endocrine therapy in the same episode | 122 | 54 | 160 |
| Excluded by QC: endocrine therapy in HR-negative disease | 9 | 0 | 16 |
| Retained episodes (patients) | 749 (270) | 381 (130) | 1,040 (295) |
| For comparison: non-LHRH BPC regimens with PFS labels in the same patients | 1,493 | 750 | 1,896 |

**b. Agreement with conditional denominators**

| BPC site | Same regimen / all retained | Same event indicator, given same regimen | Same event indicator and exact PFS duration, given same regimen | Cohen's kappa |
|---|---|---|---|---|
| DFCI | 646/749 (86.2%) | 602/646 (93.2%) | 519/646 (80.3%) | 0.600 |
| VICC | 314/381 (82.4%) | 297/314 (94.6%) | 265/314 (84.4%) | 0.595 |
| MSK | 868/1,040 (83.5%) | 827/868 (95.3%) | 658/868 (75.8%) | 0.724 |

**c. Regimen disagreement categories**

| | DFCI (749 episodes) | VICC (381 episodes) | MSK (1,040 episodes) |
|---|---|---|---|
| Same regimen | 646 (86.2%) | 314 (82.4%) | 868 (83.5%) |
| Different: treatment changed without counted progression | 60 (8.0%) | 45 (11.8%) | 88 (8.5%) |
| Different: investigational drug masked in BPC | 0 (0.0%) | 0 (0.0%) | 40 (3.8%) |
| Different: other recording differences | 39 (5.2%) | 20 (5.2%) | 32 (3.1%) |
| Different: not attributed | 4 (0.5%) | 2 (0.5%) | 12 (1.2%) |
| Same regimen or a listed grouping difference (rows 1–3) | 706 (94.3%) | 359 (94.2%) | 996 (95.8%) |

#### **Supplementary Table 19** (continued).

**d. Endpoint disagreement categories**

| | **DFCI (646 pairs)** | **VICC (314 pairs)** | **MSK (868 pairs)** |
|---|---|---|---|
| Same event indicator and exact PFS duration | 519 (80.3%) | 265 (84.4%) | 658 (75.8%) |
| BPC event is a progression report in the first 28 days; our rule does not count it | 66 (10.2%) | 26 (8.3%) | 133 (15.3%) |
| Progression reported within 28 days after a regimen switch; BPC censors at the switch | 19 (2.9%) | 7 (2.2%) | 28 (3.2%) |
| Both censored; censoring-date rules differ | 19 (2.9%) | 9 (2.9%) | 33 (3.8%) |
| Start dates differ | 1 (0.2%) | 0 (0.0%) | 5 (0.6%) |
| Other | 22 (3.4%) | 7 (2.2%) | 11 (1.3%) |
| Exact agreement or a listed endpoint-rule difference (rows 1–4) | 623 (96.4%) | 307 (97.8%) | 852 (98.2%) |

**Supplementary Table 20**: First-line median PFS by exact regimen in MSK-CHORD and GENIE BPC, with a published benchmark. **a**, MSK-CHORD: retained first captured lines, study HR/HER2 fields. GENIE BPC: first non-GnRH metastatic regimen, curated subtype, three sites pooled, PFS-I + Mixed (Progressing or Mixed imaging, or death). Exact named drug sets; the two most frequent named regimens per subtype are shown (one for HR−/HER2+; the pooled 'Other' category is excluded). Lines equal patients; events are counted after cleaning. Kaplan–Meier median and 95% CI in days; NR, not reached. The cohorts are not matched. With late-start lines restored, the HR+/HER2+ paclitaxel + pertuzumab + trastuzumab median is 434 days (76 lines); see Supplementary Table 28. **b**, Published real-world benchmark for first-line letrozole with or without palbociclib in HR-positive/HER2-negative women (DeMichele et al.): clinician-assessed progression or death; medians in months from stabilized inverse-probability-of-treatment-weighted Kaplan–Meier estimates; same named drugs, different source population and assessment. Published values are external context, not an agreement test.

**a. Regimen medians**

| | MSK-CHORD | | | GENIE BPC PFS-I + Mixed | | |
|---|---|---|---|---|---|---|
| **Subtype and exact drug set** | **n** | **Events** | **Median (95% CI)** | **n** | **Events** | **Median (95% CI)** |
| *HR+/HER2−* | | | | | | |
| Letrozole + palbociclib | 196 | 117 | 454 (385–601) | 60 | 46 | 289 (119–373) |
| Letrozole | 137 | 84 | 382 (294–473) | 57 | 38 | 254 (112–370) |
| *HR−/HER2+* | | | | | | |
| Paclitaxel + pertuzumab + trastuzumab | 54 | 26 | 658 (317–NR) | 8 | 7 | 209 (59–289) |
| *HR−/HER2−* | | | | | | |
| Capecitabine | 76 | 69 | 94 (78–120) | 26 | 20 | 77 (62–217) |
| Paclitaxel | 53 | 48 | 91 (73–144) | 6 | 5 | 144 (17–224) |
| *HR+/HER2+* | | | | | | |
| Paclitaxel + pertuzumab + trastuzumab | 41 | 28 | 233 (174–289) | 25 | 10 | 673 (208–NR) |
| Letrozole | 13 | 11 | 231 (99–339) | 7 | 4 | 416 (4–NR) |

**b. Published real-world benchmark (DeMichele et al.)**

| **Treatment** | **Patients (unweighted)** | **Median PFS, months (95% CI)** |
|---|---|---|
| Palbociclib plus letrozole | 772 | 20.0 (17.5–21.9) |
| Letrozole alone | 658 | 11.9 (10.5–13.7) |

*Reference:* DeMichele, A. et al. Comparative effectiveness of first-line palbociclib plus letrozole versus letrozole alone for HR+/HER2− metastatic breast cancer in US real-world clinical practice. *Breast Cancer Res.* **23**, 37 (2021).

**Supplementary Table 21**: First-line and all-line endpoint sensitivities in MSK-CHORD. Rules requiring no non-progression report within 30, 60 or 90 days after a positive report; a later positive report is not required. All 2,342 first lines, all 8,791 mLoTs and the saved GBSA predictions are used without retraining; n and events are rule-specific; medians are Kaplan–Meier days with 95% CI; time-dependent C and 365-day Brier score are mean ± SD across the five outer folds.

| Progression definition | n | Events | Median PFS, days (95% CI) | Time-dependent C | Brier score at 365 days |
|---|---|---|---|---|---|
| *First captured lines (mLoT1)* | | | | | |
| Original (any progression report) | 2,342 | 1,664 | 231 (215–251) | 0.663 ± 0.006 | 0.201 ± 0.009 |
| No non-progression report within 30 days | 2,342 | 1,583 | 274 (254–291) | 0.664 ± 0.004 | 0.209 ± 0.011 |
| No non-progression report within 60 days | 2,341 | 1,510 | 311 (289–329) | 0.663 ± 0.010 | 0.220 ± 0.010 |
| No non-progression report within 90 days | 2,340 | 1,395 | 378 (353–404) | 0.661 ± 0.009 | 0.239 ± 0.013 |
| *All mLoTs* | | | | | |
| Original (any progression report) | 8,791 | 7,565 | 115 (112–118) | 0.681 ± 0.006 | 0.119 ± 0.006 |
| No non-progression report within 30 days | 8,791 | 7,354 | 136 (132–141) | 0.678 ± 0.006 | 0.132 ± 0.008 |
| No non-progression report within 60 days | 8,790 | 7,125 | 161 (156–166) | 0.677 ± 0.008 | 0.150 ± 0.009 |
| No non-progression report within 90 days | 8,789 | 6,746 | 198 (190–203) | 0.677 ± 0.007 | 0.185 ± 0.009 |

**Supplementary Table 21** (continued). Midpoint-imputation sensitivity in 2,342 first captured metastatic lines. Each imaging progression event was moved halfway between the preceding within-line non-progression report and the progression report, or halfway from line start when no earlier non-progression report was available. Death and censoring dates were unchanged. Saved GBSA predictions were evaluated without retraining.

| Measure | Original scan date | Midpoint imputation |
|---|---|---|
| Median PFS, days (95% CI) | 231 (215–251) | 177.5 (162.5–195.5) |
| Time-dependent C, mean ± fold SD | 0.663 ± 0.006 | 0.653 ± 0.007 |
| Brier score at 365 days, mean ± fold SD | 0.201 ± 0.009 | 0.195 ± 0.009 |

**Supplementary Table 22**: Repeated treatment lines. MSK-CHORD pooled out-of-fold GBSA predictions (8,791 lines, 2,881 patients). Comparable ordered pairs and C for all pairs, within-patient pairs and between-patient pairs; pair counts describe the C calculation and do not establish statistical independence of lines. Pooled C within first and later lines is reported in Supplementary Table 24c.

| Pairs included | Comparable ordered pairs | C |
|---|---|---|
| All comparable pairs (original pooled C) | 36,896,080 | 0.6807 |
| Within-patient pairs only | 15,826 | 0.6283 |
| Between-patient pairs only | 36,880,254 | 0.6807 |

**Supplementary Table 22** (continued). One randomly selected line per patient. The selection was repeated 30 times, each time choosing one line uniformly at random from each of the 2,881 patients; C is the mean (SD) over the 30 selections, using the saved pooled out-of-fold GBSA predictions without retraining. The mean difference from the pooled C over all lines is +0.02645 (95% CI +0.02105 to +0.03186), with an approximate interval from a patient-level jackknife that removes each patient's lines together while predictions stay fixed.

| **Evaluation set** | **Lines** | **C** |
|---|---|---|
| All modeled lines (original pooled C) | 8,791 | 0.6807 |
| One random line per patient, mean (SD) of 30 selections | 2,881 | 0.7071 (0.0034) |

**Supplementary Table 23**: Clinical Cox reference and full multimodal models. Results use pooled out-of-fold predictions for 8,791 mLoTs from 2,881 patients. The six-variable reference uses first-line status, HR status, HER2 status, most recent ECOG, liver involvement in the preceding 90 days, and most recent log-transformed CA15–3, with Cox regularization $\alpha = 1$. Confidence intervals use 1,000 patient-level bootstrap resamples, retaining all lines of each sampled patient. Paired comparisons use the same patient draws for both models. Predictions remain fixed, so intervals do not include model-training uncertainty.

**a. Pooled out-of-fold performance**

| **Model** | **C (95% CI)** | **IBS** |
|---|---|---|
| Six-variable clinical Cox | 0.6426<br>[0.6357, 0.6502] | 0.1343 |
| Full CoxPH | 0.6782<br>[0.6719, 0.6846] | 0.1257 |
| Full GBSA | 0.6807<br>[0.6741, 0.6873] | 0.1241 |

**b. Paired differences: full model minus clinical reference**

| **Full model** | $\Delta C$ **(paired 95% CI)** | **ΔIBS (paired 95% CI)** |
|---|---|---|
| CoxPH | +0.0355<br>[0.0305, 0.0405] | −0.008551<br>[−0.009933, −0.007214] |
| GBSA | +0.0380<br>[0.0331, 0.0429] | −0.010140<br>[−0.011578, −0.008754] |

*Notes:* Higher C and lower IBS indicate better performance. Relative gain in discrimination above chance is $(C_{\text{full}} - C_{\text{reference}})/(C_{\text{reference}} - 0.5)$: approximately 25% for full CoxPH and 27% for full GBSA. These percentages describe the increase in $C - 0.5$, not the relative increase in C itself or clinical benefit. For paired IBS intervals, the censoring estimate and observed censoring-time support are recalculated within each patient draw. Within each outer fold, $\Delta C$ ranged from 0.0303 to 0.0458 for full CoxPH and from 0.0320 to 0.0468 for full GBSA, favoring the full model in all five folds.

**Supplementary Table 24**: Sensitivity to first-line status and missingness indicators, and discrimination within line groups. MSK-CHORD pooled out-of-fold GBSA predictions, 8,791 mLoTs from 2,881 patients. **a**, Full GBSA and GBSA without first-line status and 14 explicit missingness or unknown indicators (15 inputs removed); other recorded care-process information remains. **b**, Paired change (reduced minus full model). **c**, Pooled C of the full GBSA within first lines (mLoT1) and later lines (mLoT2+); group-specific C is conditional on line group. Intervals are 95% percentile intervals from 1,000 patient resamples that retain all lines of each sampled patient, with predictions fixed; paired intervals use the same resamples for both models.

**a. Pooled out-of-fold performance**

| **Model** | **C (95% CI)** | **IBS** |
|---|---|---|
| Full GBSA | 0.6807<br>[0.6741, 0.6873] | 0.1241 |
| GBSA without first-line status and missingness indicators | 0.6784<br>[0.6720, 0.6852] | 0.1249 |

**b. Paired difference: reduced minus full GBSA**

| **Comparison** | $\Delta C$<br>**(paired 95% CI)** | **ΔIBS**<br>**(paired 95% CI)** |
|---|---|---|
| Reduced minus full GBSA | −0.002254<br>[−0.003621, −0.000835] | +0.000785<br>[+0.000341, +0.001230] |

**c. Full GBSA within line groups**

| **Line group** | **Lines (patients)** | **C (95% CI)** |
|---|---|---|
| All lines | 8,791 (2,881) | 0.6807<br>[0.6741, 0.6873] |
| First lines (mLoT1) | 2,342 (2,342) | 0.6618<br>[0.6483, 0.6755] |
| Later lines (mLoT2+) | 6,449 (2,149) | 0.6568<br>[0.6488, 0.6648] |

*Notes:* Higher C and lower IBS indicate better performance. For the paired IBS interval, the censoring estimate and observed censoring-time support are recalculated within each patient draw.

**Supplementary Table 25**: Internal and external calibration at 180, 365 and 730 days. Point estimates with 95% percentile intervals from 1,000 patient resamples that retain all lines of each sampled patient; censoring estimates and calibration models are refitted in each resample while prediction models and spline knots stay fixed, so intervals do not include model-training uncertainty. CITL (calibration-in-the-large) is on the log-odds scale (ideal 0; positive values indicate average under-estimation of progression risk); the ideal calibration slope is 1 (values above 1 indicate compressed predictions); ICI, E50 and E90 are the mean, median and 90th percentile of absolute calibration error from an IPCW three-knot natural cubic-spline calibration curve, in percentage points. **a**, MSK-CHORD out-of-fold predictions. **b**, DFCI and **c**, VICC, BPC PFS-I + Mixed with the 28-day minimum and 730-day cap reapplied; external probabilities average the five fixed MSK-CHORD fold models. **d**, DFCI and **e**, VICC, before and after a patient-level cross-validated intercept-and-slope update; resampling details are given with these panels. **f**, DFCI and VICC regimens pooled: ICI from the same calibration diagnostic fitted with pooled censoring weights to the original and to the site-specific cross-validated updated probabilities; point estimates only.

**a. MSK-CHORD, out-of-fold predictions (8,791 lines, 2,881 patients)**

| Measure | 180 days | 365 days | 730 days |
|---|---|---|---|
| CITL | −0.042 [−0.092, 0.009] | −0.085 [−0.150, −0.014] | −0.128 [−0.217, −0.034] |
| Calibration slope | 0.986 [0.930, 1.046] | 1.078 [0.998, 1.156] | 1.000 [0.903, 1.106] |
| ICI (percentage points) | 1.75 [1.16, 2.49] | 1.57 [1.04, 2.23] | 0.91 [0.38, 1.59] |
| E50 (percentage points) | 1.57 [0.87, 2.51] | 0.65 [0.34, 1.33] | 0.32 [0.10, 0.97] |
| E90 (percentage points) | 3.45 [2.28, 5.39] | 5.22 [2.86, 7.42] | 3.01 [0.84, 5.32] |
| **b. DFCI, BPC PFS-I + Mixed (1,394 regimens, 317 patients)** | | | |
| CITL | 0.485 [0.316, 0.650] | 0.237 [0.057, 0.449] | 0.090 [−0.134, 0.376] |
| Calibration slope | 1.470 [1.178, 1.807] | 1.426 [1.153, 1.744] | 1.371 [1.024, 1.836] |
| ICI (percentage points) | 11.28 [7.80, 14.87] | 4.65 [2.55, 8.01] | 1.72 [1.02, 4.72] |
| E50 (percentage points) | 12.64 [7.79, 16.28] | 4.89 [1.82, 8.75] | 1.46 [0.85, 4.90] |
| E90 (percentage points) | 15.85 [12.09, 20.88] | 7.29 [4.74, 11.16] | 3.48 [1.77, 8.48] |
| **c. VICC, BPC PFS-I + Mixed (695 regimens, 152 patients)** | | | |
| CITL | 0.555 [0.356, 0.746] | 0.462 [0.225, 0.784] | 0.430 [0.016, 0.920] |
| Calibration slope | 1.484 [1.062, 2.047] | 1.678 [1.099, 2.461] | 1.591 [0.877, 2.688] |
| ICI (percentage points) | 12.94 [8.92, 17.10] | 9.16 [5.78, 13.27] | 5.26 [2.39, 8.87] |
| E50 (percentage points) | 14.37 [9.22, 18.87] | 10.20 [5.93, 14.81] | 5.88 [2.18, 9.79] |
| E90 (percentage points) | 17.58 [11.99, 24.05] | 13.02 [8.16, 18.80] | 7.72 [3.62, 14.42] |

**Supplementary Table 25** (continued).

**d. DFCI: before and after recalibration**

| Measure | Predictions | 180 days | 365 days | 730 days |
|---|---|---|---|---|
| ICI | Original | 11.28 [7.80, 14.87] | 4.65 [2.55, 8.01] | 1.72 [1.02, 4.72] |
| | Updated | 0.79 [0.66, 4.76] | 0.71 [0.60, 4.12] | 1.07 [0.59, 4.15] |
| CITL | Original | 0.485 [0.316, 0.650] | 0.237 [0.057, 0.449] | 0.090 [−0.134, 0.376] |
| | Updated | −0.002 [−0.183, 0.174] | −0.003 [−0.196, 0.221] | 0.000 [−0.237, 0.296] |
| Calibration slope | Original | 1.470 [1.178, 1.807] | 1.426 [1.153, 1.744] | 1.371 [1.024, 1.836] |
| | Updated | 0.950 [0.761, 1.179] | 0.950 [0.759, 1.164] | 0.949 [0.700, 1.280] |
| E50 | Original | 12.64 [7.79, 16.28] | 4.89 [1.82, 8.75] | 1.46 [0.85, 4.90] |
| | Updated | 0.74 [0.46, 5.07] | 0.64 [0.44, 4.45] | 0.70 [0.31, 3.90] |
| E90 | Original | 15.85 [12.09, 20.88] | 7.29 [4.74, 11.16] | 3.48 [1.77, 8.48] |
| | Updated | 1.70 [1.22, 7.99] | 1.21 [1.05, 7.67] | 1.59 [1.02, 8.11] |

**e. VICC: before and after recalibration**

| Measure | Predictions | 180 days | 365 days | 730 days |
|---|---|---|---|---|
| ICI | Original | 12.94 [8.92, 17.10] | 9.16 [5.78, 13.27] | 5.26 [2.39, 8.87] |
| | Updated | 0.71 [0.84, 5.69] | 1.81 [1.03, 5.65] | 0.97 [0.83, 5.36] |
| CITL | Original | 0.555 [0.356, 0.746] | 0.462 [0.225, 0.784] | 0.430 [0.016, 0.920] |
| | Updated | −0.000 [−0.215, 0.199] | −0.002 [−0.265, 0.338] | −0.006 [−0.462, 0.514] |
| Calibration slope | Original | 1.484 [1.062, 2.047] | 1.678 [1.099, 2.461] | 1.591 [0.877, 2.688] |
| | Updated | 0.962 [0.683, 1.332] | 0.912 [0.562, 1.397] | 0.803 [0.364, 1.426] |
| E50 | Original | 14.37 [9.22, 18.87] | 10.20 [5.93, 14.81] | 5.88 [2.18, 9.79] |
| | Updated | 0.69 [0.62, 5.81] | 1.95 [0.77, 5.97] | 1.04 [0.53, 4.96] |
| E90 | Original | 17.58 [11.99, 24.05] | 13.02 [8.16, 18.80] | 7.72 [3.62, 14.42] |
| | Updated | 1.46 [1.45, 10.76] | 2.96 [1.69, 10.98] | 1.59 [1.35, 10.80] |

**f. DFCI and VICC pooled (2,089 regimens, 469 patients): ICI before and after the site-specific updates**

| Measure | Predictions | 180 days | 365 days | 730 days |
|---|---|---|---|---|
| ICI | Original | 11.86 | 6.05 | 2.62 |
| | Updated | 0.80 | 0.51 | 0.59 |

*Notes:* ICI is the primary calibration measure. ICI, E50 and E90 are in percentage points; CITL and calibration slope are unscaled. Original probabilities average the five fixed GBSA models. Updated probabilities use a horizon-specific intercept-and-slope update evaluated with five patient-level folds within each site. Each regimen receives an update fitted on the other four folds. The GBSA models remain fixed. Values are point estimates with 95% percentile intervals from 1,000 patient resamples. Updated probabilities remain fixed within resamples; the update is not refitted. For absolute-error measures near zero, percentile intervals can lie above the point estimate. Panel f pools the DFCI and VICC regimens and fits the calibration diagnostic with pooled censoring weights; it is a pooled diagnostic, not an average of the site ICIs, and no resampling interval is reported for it.

**Supplementary Table 26**: Sensitivity to the number of recorded metastatic-site categories. MSK-CHORD pooled out-of-fold predictions, 8,791 mLoTs from 2,881 patients. The added GBSA input counts the recorded metastatic-site categories (of ten) in the 90 days before line start; it counts categories, not lesions or disease volume. **a**, Pooled out-of-fold performance. **b**, Paired change, site-count model minus original GBSA. Intervals use 1,000 paired patient resamples with all lines of each sampled patient retained and the fitted models fixed, so they do not include model-training uncertainty.

**a. Pooled out-of-fold performance**

| Model | C (95% CI) | IBS |
|---|---|---|
| Original GBSA | 0.68065<br>[0.67405, 0.68731] | 0.12412 |
| GBSA plus site count | 0.68119<br>[0.67453, 0.68767] | 0.12405 |

**b. Paired differences: site count minus original**

| Comparison | $\Delta C$<br>(paired 95% CI) | ΔIBS<br>(paired 95% CI) |
|---|---|---|
| Site count minus original | +0.00054<br>[−0.00064, +0.00174] | −0.00007<br>[−0.00045, +0.00028] |

*Notes:* Higher C and lower IBS indicate better performance. For the paired IBS interval, the censoring estimate and observed censoring-time support are recalculated within each patient draw.

**Supplementary Table 27**: Sensitivity to the pre-line imaging requirement. MSK-CHORD saved GBSA predictions after relaxing pre-line imaging eligibility from 90 to 180 days with predictor windows and QC unchanged; 1,000 paired patient resamples from the 2,922-patient union, models fixed; the original-cohort interval resamples its own 2,881 patients, as in Supplementary Table 23; IBS is descriptive; of the 244 patients in the added group, 41 were absent from the original cohort and therefore from all original training folds.

| Cohort | Lines | Patients | C (95% CI) | IBS |
|---|---|---|---|---|
| Original 90-day cohort | 8,791 | 2,881 | 0.6807 [0.6741, 0.6873] | 0.1241 |
| Expanded 180-day cohort | 9,055 | 2,922 | 0.6828 [0.6758, 0.6891] | 0.1265 |
| Added 91–180-day group | 264 | 244 | 0.6638 [0.6210, 0.7093] | 0.2073 |
| Paired change, expanded minus original | | | +0.0022 [0.0007, 0.0037] | |

**Supplementary Table 28**: Late-start chemotherapy plus endocrine QC exclusions in MSK-CHORD. Lines flagged by the chemotherapy plus endocrine QC rule are grouped by the treatment classes recorded within the first 28 days after line start, the model's treatment-input window; in late-start lines, at least one class first appeared after day 28. Of the 525 late-start lines, 20 also failed the HR-negative endocrine rule; the other 505 are the restored lines. PFS uses the pre-QC labels with the 730-day cap; medians are Kaplan–Meier estimates. **a**, Lines flagged by the rule. **b**, GBSA refits on the original five patient-level outer folds with the saved hyperparameters; restored lines of existing patients follow their original fold, new patients were assigned to folds with a fixed seed, and later agents remain masked from the model inputs. C is pooled adjusted-Antolini concordance on the same 8,791 original lines for every model; paired intervals use 1,000 patient resamples with predictions fixed. **c**, One-year calibration of the original models and the refit with the 505 late-start lines restored (full PFS), in the 9,296-line cohort against the same full-PFS labels; original-model predictions for restored lines use a saved fold model that had not trained on that patient, and refit predictions are out of fold. **d**, The same two sets of fold models (five-model average) on GENIE BPC PFS-I + Mixed, before recalibration. **e**, Reconstruction agreement with GENIE BPC regimens, as in Supplementary Table 19, for retained episodes and for all formed episodes meeting the label, imaging and minimum-duration requirements with the QC filter removed. **f**, BPC episodes excluded as late-start by the chemotherapy plus endocrine rule: whether the later class is recorded in a different BPC regimen from the courses present in the first 28 days, and same-regimen matches using all drugs of the episode or only the drugs started by day 28, with the same one-to-one rule and 14-day start tolerance. **g**, First-line median PFS in the retained cohort and with the 505 late-start lines restored; exact initial drug sets (agents started within 28 days of line start), whose retained rows match Supplementary Table 20.

**a. Lines flagged by the chemotherapy plus endocrine rule**

| Group | Lines | Patients | First lines | Events (%) | Median PFS (days) |
|---|---|---|---|---|---|
| Chemotherapy start; endocrine therapy added later | 278 | 269 | 174 | 54.7 | 482 |
| Endocrine start; chemotherapy added later | 244 | 235 | 69 | 81.6 | 269 |
| Neither class in the first 28 days; both added later | 3 | 3 | 0 | 66.7 | 402 |
| Late-start lines, total | 525 | 489 | 243 | 67.2 | 344 |
| Both classes recorded within the first 28 days | 1,589 | 985 | 244 | 91.3 | 113 |
| Original retained cohort | 8,791 | 2,881 | 2,342 | 86.1 | 115 |

**b. Refits: discrimination on the original 8,791 lines**

| Refit cohort | Lines | C on original 8,791 lines | Paired ΔC (95% CI) | Paired ΔIBS (95% CI) |
|---|---|---|---|---|
| Original analysis | 8,791 | 0.6807 | reference | reference |
| 505 late-start lines restored, full PFS | 9,296 | 0.6799 | −0.0008 [−0.0017, +0.0001] | −0.0001 [−0.0005, +0.0003] |
| 505 late-start lines censored at addition | 9,296 | 0.6802 | −0.0004 [−0.0013, +0.0004] | +0.0000 [−0.0003, +0.0004] |
| Late-start and HR-positive GnRH groups restored | 9,485 | 0.6800 | −0.0007 [−0.0017, +0.0003] | −0.0001 [−0.0005, +0.0003] |
| Chemotherapy plus endocrine rule removed | 10,824 | 0.6808 | +0.0002 [−0.0010, +0.0013] | −0.0001 [−0.0005, +0.0003] |
| All QC exclusions restored | 11,085 | 0.6810 | +0.0003 [−0.0009, +0.0015] | −0.0002 [−0.0007, +0.0002] |

**Supplementary Table 28** (continued).

**c. One-year calibration in the 9,296-line cohort, original and refit models**

| Stratum | Lines | Observed one-year PFS, % (95% CI) | Model | Mean predicted, % | Predicted − observed, points | Slope | ICI, points |
|---|---|---|---|---|---|---|---|
| HR+/HER2+ first lines | 159 | 55.1 [46.6, 62.9] | Original | 38.2 | −17.0 | 0.853 | 17.83 |
| starting on chemotherapy | | | Refit | 44.6 | −10.5 | 1.138 | 11.93 |
| HR+/HER2− first lines | 373 | 37.0 [31.9, 42.1] | Original | 27.3 | −9.7 | 1.705 | 11.02 |
| starting on chemotherapy | | | Refit | 33.6 | −3.4 | 1.918 | 10.70 |
| All HR+/HER2+ | 315 | 45.4 [39.6, 51.1] | Original | 39.0 | −6.5 | 0.907 | 7.05 |
| first lines | | | Refit | 43.8 | −1.6 | 1.200 | 3.14 |
| HR+ endocrine-start lines | 3,836 | 29.6 [28.1, 31.1] | Original | 26.5 | −3.1 | 1.177 | 3.45 |
| including restored | | | Refit | 27.7 | −1.9 | 1.171 | 2.52 |
| Triple-negative | 1,032 | 10.9 [9.0, 13.0] | Original | 11.6 | +0.7 | 0.844 | 1.28 |
| | | | Refit | 11.9 | +1.0 | 0.810 | 1.67 |
| HR−/HER2+ | 437 | 28.6 [24.2, 33.0] | Original | 22.9 | −5.6 | 0.995 | 5.61 |
| | | | Refit | 23.4 | −5.2 | 1.004 | 5.28 |
| All lines | 9,296 | 20.2 [19.4, 21.1] | Original | 18.1 | −2.2 | 1.096 | 2.24 |
| | | | Refit | 19.2 | −1.1 | 1.100 | 1.81 |
| Restored late-start lines | 505 | 47.1 [42.5, 51.4] | Original | 25.0 | −22.0 | 1.230 | 22.25 |
| only | | | Refit | 29.1 | −17.9 | 1.240 | 18.24 |

**d. External evaluation before recalibration, original / refit**

| Site | Regimens | Patients | C | IBS | 180-day ICI, points |
|---|---|---|---|---|---|
| DFCI | 1,394 | 317 | 0.6508 / 0.6542 | 0.1656 / 0.1682 | 11.28 / 13.36 |
| VICC | 695 | 152 | 0.6352 / 0.6363 | 0.1589 / 0.1627 | 12.94 / 15.00 |

**e. Reconstruction agreement with GENIE BPC before QC**

| | Same regimen | | Same event, given same regimen | |
|---|---|---|---|---|
| Site | Retained episodes | QC filter removed | Retained episodes | QC filter removed |
| DFCI | 646/749 (86.2%) | 718/880 (81.6%) | 602/646 (93.2%) | 669/718 (93.2%) |
| VICC | 314/381 (82.4%) | 334/435 (76.8%) | 297/314 (94.6%) | 315/334 (94.3%) |
| MSK | 868/1,040 (83.5%) | 944/1,216 (77.6%) | 827/868 (95.3%) | 897/944 (95.0%) |

**f. Late-start BPC episodes excluded by the chemotherapy plus endocrine rule**

| Site | Episodes | Later class in a separate BPC regimen | Same regimen, all drugs | Same regimen, drugs started by day 28 |
|---|---|---|---|---|
| DFCI | 37 | 37/37 (100.0%) | 0/37 (0.0%) | 27/37 (73.0%) |
| VICC | 21 | 21/21 (100.0%) | 1/21 (4.8%) | 17/21 (81.0%) |
| MSK | 69 | 69/69 (100.0%) | 1/69 (1.4%) | 54/69 (78.3%) |
| All sites | 127 | 127/127 (100.0%) | 2/127 (1.6%) | 98/127 (77.2%) |

**g. First-line median PFS with late-start lines restored**

| | Retained cohort | | Late-start lines restored | |
|---|---|---|---|---|
| Subtype and exact initial drug set | Lines | Median (days) | Lines | Median (days) |
| HR+/HER2−: all first lines | 1,565 | 275 | 1,730 | 301 |
| Letrozole + palbociclib | 196 | 454 | 203 | 448 |
| Letrozole | 137 | 382 | 146 | 385 |
| HR+/HER2+: all first lines | 247 | 240 | 315 | 310 |
| Paclitaxel + pertuzumab + trastuzumab | 41 | 233 | 76 | 434 |

*Notes:* Slope is the IPCW logistic calibration slope (ideal 1); ICI is in percentage points, as in Supplementary Table 25. Predicted − observed is mean predicted minus observed one-year PFS, computed before rounding; negative values indicate under-prediction of PFS. Restored late-start lines are defined by a treatment event after line start and are shown separately from the strata defined by subtype and initial treatment. No late-start lines were restored in HR−/HER2+ or triple-negative disease.

### Figures

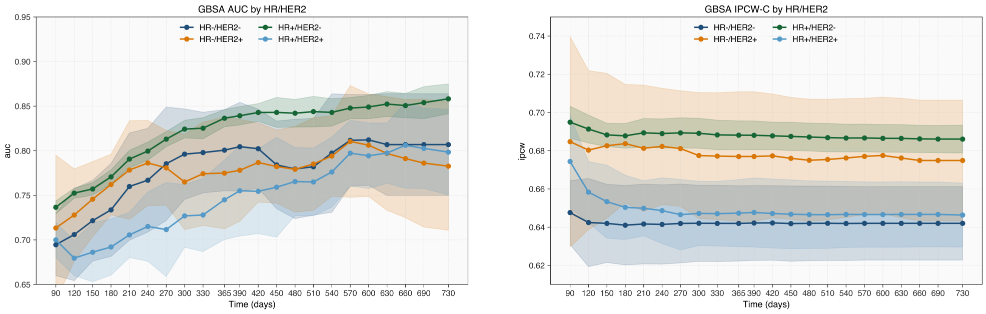


(a) Time-dependent AUC by subtype.

(b) Time-dependent IPCW C-index by subtype.

**Supplementary Figure 1**: Subtype-stratified time-dependent discrimination of the gradient-boosted Cox survival model (GBSA) for PFS prediction. **a** Cumulative/dynamic AUC and **b** inverse-probability-of-censoring-weighted (IPCW) C-index over time, shown as mean with shaded ±1 SD across the five outer cross-validation folds. HR, hormone receptor; HER2, human epidermal growth factor receptor 2.

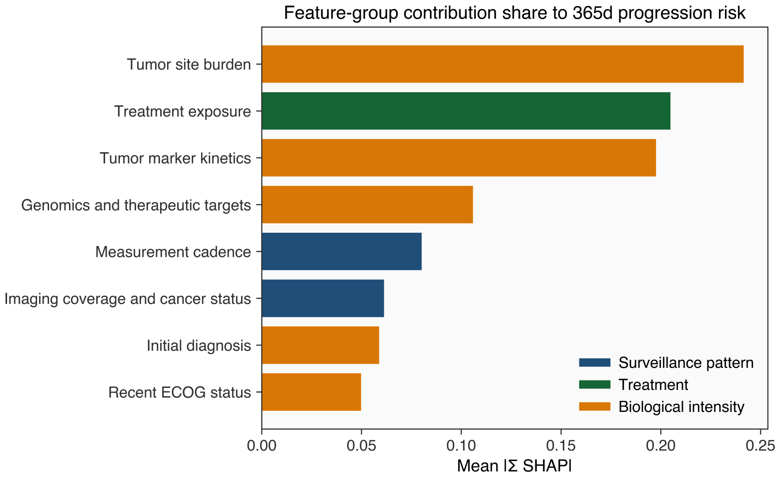


**Supplementary Figure 2**: Mean normalized SHAP contributions aggregated by clinical feature groups (bars colored by surveillance-pattern, treatment, and biological-intensity domains).

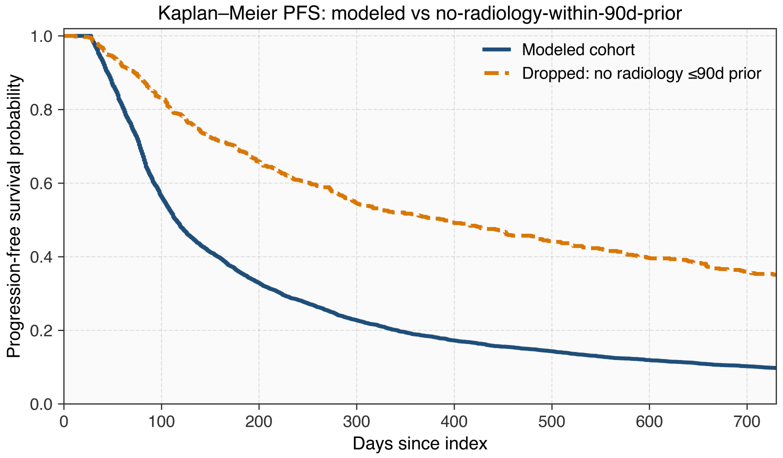


**Supplementary Figure 3**: Kaplan–Meier estimates of PFS for modeled mLoTs compared with lines excluded for missing radiology screening within 90 d before line start.

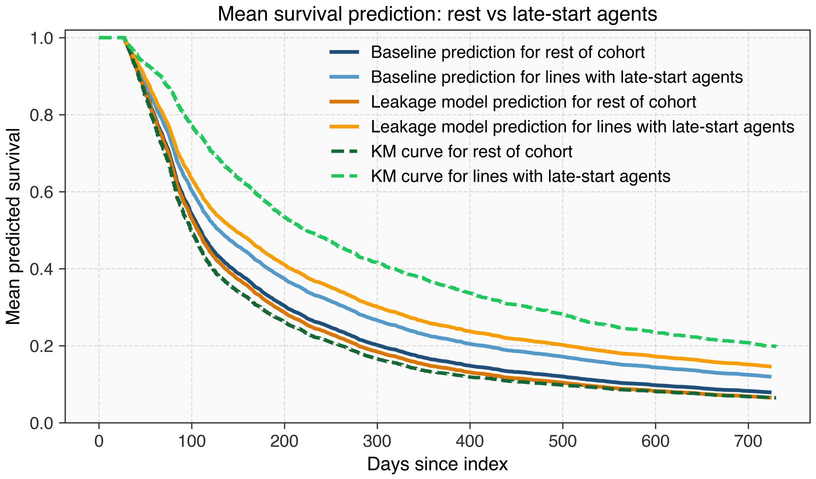


**Supplementary Figure 4**: Late-start treatment leakage analysis. Observed and predicted survival for lines in which all therapies are initiated within 28 days of line start versus lines that include late-start therapies. Dashed lines indicate Kaplan–Meier estimates of PFS; solid lines show mean predicted survival from a standard treatment-masked model (blue) and a model trained without treatment masking (orange), included here as a leakage demonstration rather than a clinical candidate model.

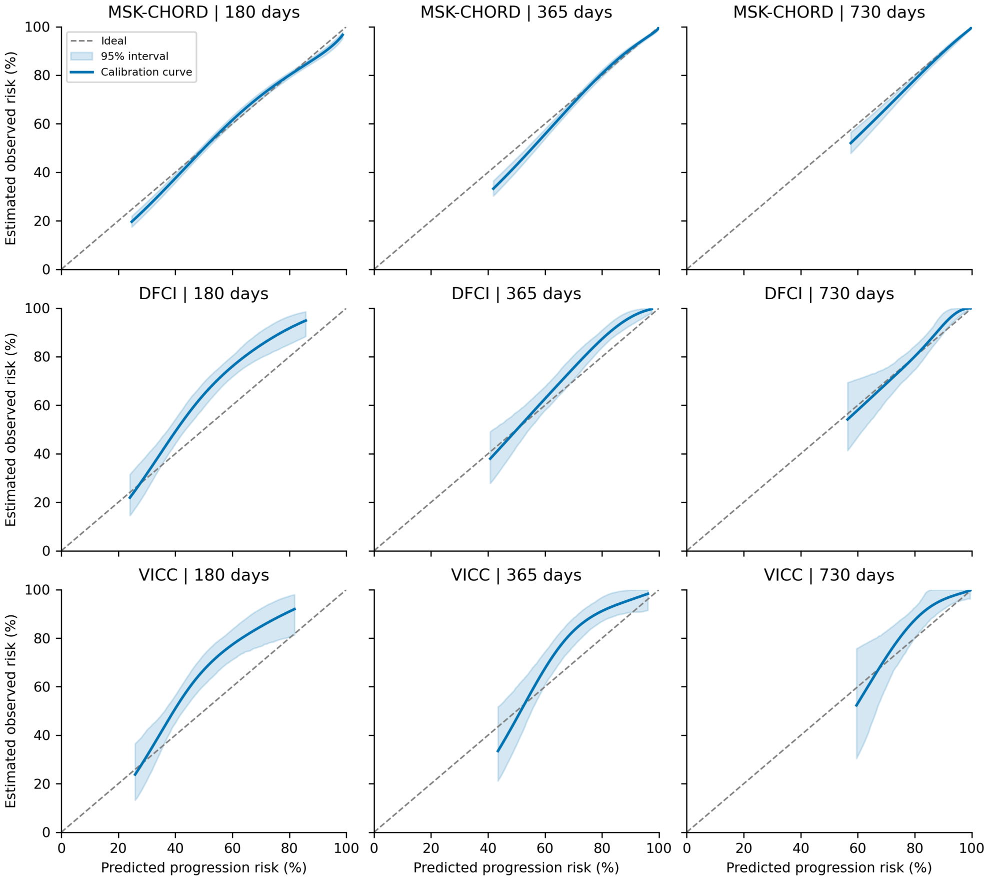


**Supplementary Figure 5**: Calibration curves at 180, 365 and 730 days. Rows show MSK-CHORD (8,791 lines, 2,881 patients), DFCI (1,394 regimens, 317 patients) and VICC (695 regimens, 152 patients). Curves estimate observed progression risk against predicted risk using IPCW and the three-knot natural cubic spline. Shading shows pointwise 95% intervals from 1,000 patient resamples with prediction models and spline knots fixed. The dashed line indicates ideal calibration. Curves are displayed over the 1st–99th percentiles of predicted risk. Internal predictions are out-of-fold; external predictions average the five fixed GBSA models before local recalibration.

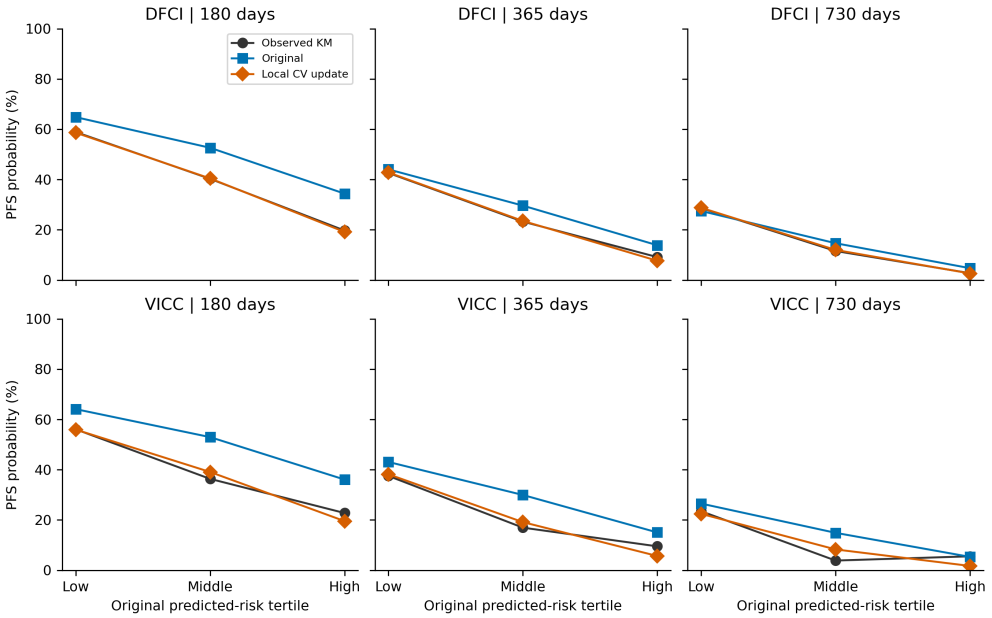


**Supplementary Figure 6**: Observed and predicted PFS by original predicted-risk tertile at DFCI and VICC. Tertiles use each site's original predicted progression risk at 365 days. At each horizon, points show Kaplan–Meier survival and mean predicted survival before and after the site-specific, patient-level cross-validated intercept-and-slope update. Each regimen's update was fitted on patients in the other four folds. GBSA models remained fixed. Curves connect descriptive point estimates; no intervals are shown. The DFCI tertiles contain 465, 464 and 465 regimens; the VICC tertiles contain 232, 231 and 232 regimens. Patients can contribute lines to more than one tertile. Two-year follow-up is limited, with 101 DFCI and 29 VICC regimens observed event-free through 730 days.

#### Model parameters and feature variables

##### Supplementary Note 1: Feature variables

The complete list of 205 model inputs and their modality assignments is provided in Supplementary Table 18. The table also identifies the 29 overlapping surveillance-pattern inputs and each retained gene indicator.

***Clinicopathologic*** Static demographics and baseline diagnostic/pathology summaries at line initiation.

- Demographics/receptors: age, sex, HR, HER2.
- Receptor status: HR and HER2 use static patient-level MSK-CHORD fields across lines; the pipeline does not select a new receptor assessment at each line start. These fields do not identify a selected primary or metastatic assay. Changes in receptor status can therefore cause subtype misclassification.
- Diagnosis/pathology: stage at diagnosis (CDM-derived), clinical/pathology groupings, histology, summary stage, and primary breast subsite categories.
- ECOG: most recent value (last 90d), days since last ECOG, missingness, and an indicator for any ECOG $\geq 2$ within 180d.

***Tumor markers*** Longitudinal CA15–3 and CEA labs in retrospective windows $T \in \{60, 180\}$ days before line start.

- Per window: exp.-weighted mean (log), max/min (log), slope (log vs time), change vs prior (log).
- Additional indicators: last observed value (log), days since last observation, missingness, and threshold/rise flags.

***Radiology*** Disease status derived from NLP-labeled radiology reports.

- Tumor sites: organ-site tumor presence within the last 90 days across 10 sites, additionally encoded for presence ever prior to line start.
- Imaging coverage/cancer status: for each anatomical region (head, chest, abdomen, pelvis, other), indicators for the associated cancer status (present/absent/indeterminate/no radiology report available) mentioned in the report in which the region is screened, plus an ever-positive indicator for whether cancer was ever reported when that region was screened prior to line start. Several body parts could be screened in the same report and share the same cancer status indicator.

***Treatment*** Planned regimen context at line initiation.

- Planned agents: therapies initiated within 28 days of line start or active at line start (carryover by stop dates).
- Encoding: within each therapy class (endocrine, targeted, biologic, chemotherapy, immunotherapy, bone-directed, other), one-hot the top 5 agents and bucket rarer agents as *other*.
- Local therapy: surgery and radiation therapy within 28 days of line start.

***Genomics*** Curated somatic alterations from tumor sequencing.

- PD-L1 and MMR: encoded as {positive, negative, unknown} due to sparse testing.
- The retained gene indicators are mutations (25): AKT1, ARID1A, ATM, BRCA1, BRCA2, CBFB, CDH1, CDKN1B, ESR1, FOXA1, GATA3, KMT2C, KMT2D, MAP2K4, MAP3K1, NCOR1, NF1, PIK3CA, PIK3R1, PTEN, RB1, RUNX1, SF3B1, TBX3, and TP53; amplifications (12): CCND1, CCNE1, EGFR, ERBB2, FGFR1, FGFR2, FOXA1, IGF1R, MCL1, MYC, PAK1, and PIK3CA; deletions (8): ATM, BRCA1, BRCA2, CDKN2A, CDKN2B, MAP2K4, PTEN, and RB1. Mutations use the source mutation records; amplification and deletion indicators use source CNA calls of $+2$ and $-2$, respectively.
- Timing and missingness: gene indicators are set to zero and `GENOMICS_MISSING` is set to one for lines before the sequenced specimen date, or when that date is unavailable. This is a line-level flag.

#### Supplementary Note 2: Model parameters

- CoxPH (regularized): L2 penalty strength $\alpha$ tuned over 30 log-spaced values between $10^{-1}$ and $10^{3}$ (rounded to 3 decimals).
- Gradient boosted survival trees (CoxPH loss; GBSA) configurations:
  - gbsa-simple-400: n_estimators=400, learning_rate=0.05, max_depth=2, min_samples_split=10, min_samples_leaf=50, subsample=0.5, max_features=0.3.
  - gbsa-mid-400: n_estimators=400, learning_rate=0.05, max_depth=3, min_samples_split=10, min_samples_leaf=50, subsample=0.7, max_features=0.3.
  - gbsa-large-400: n_estimators=400, learning_rate=0.05, max_depth=3, min_samples_split=10, min_samples_leaf=5, subsample=1.0, max_features=0.5.
  - gbsa-simple-800: n_estimators=800, learning_rate=0.05, max_depth=2, min_samples_split=10, min_samples_leaf=50, subsample=0.5, max_features=0.3.
  - gbsa-mid-800: n_estimators=800, learning_rate=0.05, max_depth=3, min_samples_split=10, min_samples_leaf=50, subsample=0.7, max_features=0.3.
- Random survival forest (RSF):
  - rsf-simple-400: n_estimators=400, min_samples_split=10, min_samples_leaf=50, max_features=0.3.
  - rsf-simple-800: n_estimators=800, min_samples_split=10, min_samples_leaf=50, max_features=0.3.
  - rsf-mid-400: n_estimators=400, min_samples_split=10, min_samples_leaf=20, max_features=0.3.
  - rsf-mid-800: n_estimators=800, min_samples_split=10, min_samples_leaf=20, max_features=0.3.
  - rsf-large-400: n_estimators=400, min_samples_split=10, min_samples_leaf=5, max_features=0.5.
- DeepSurv: MLP trained for max num_epochs=200 with batch_size=256. A subset of 10% of the available training data for each fold (outer and inner) is held out for early stopping.
  - ds-lite-64-strongreg: hidden_sizes=[64], dropout=0.4, lr=$2 \times 10^{-4}$, weight_decay=$1 \times 10^{-3}$.
  - ds-lite-64-weakreg: hidden_sizes=[64], dropout=0.2, lr=$2 \times 10^{-4}$, weight_decay=$5 \times 10^{-4}$.
  - ds-base-128-strongreg: hidden_sizes=[128], dropout=0.4, lr=$3 \times 10^{-4}$, weight_decay=$1 \times 10^{-3}$.
  - ds-base-128-weakreg: hidden_sizes=[128], dropout=0.2, lr=$3 \times 10^{-4}$, weight_decay=$5 \times 10^{-4}$.

- ds-mlp-128x32-strongreg: hidden_sizes=[128, 32], dropout=0.4, lr=$5\times10^{-4}$, weight_decay=$1\times10^{-3}$.

- DeepHit: discretized time grid with num_time_steps $\in \{104, 196\}$; trained for max num_epochs=200 with batch_size=256 and fixed alpha=0.5. A subset of 10% of the available training data for each fold (outer and inner) is held out for early stopping.
  - dh104-base-strongreg: num_time_steps=104, hidden_dims=[128], dropout=0.4, lr=$2\times10^{-4}$, weight_decay=$1\times10^{-3}$.
  - dh104-base-weakreg: num_time_steps=104, hidden_dims=[128], dropout=0.2, lr=$2\times10^{-4}$, weight_decay=$5\times10^{-4}$.
  - dh104-mid-strongreg: num_time_steps=104, hidden_dims=[64, 32], dropout=0.4, lr=$5\times10^{-4}$, weight_decay=$1\times10^{-3}$.
  - dh104-mid-weakreg: num_time_steps=104, hidden_dims=[64, 32], dropout=0.2, lr=$5\times10^{-4}$, weight_decay=$5\times10^{-4}$.
  - dh196-large-strongreg: num_time_steps=196, hidden_dims=[128, 64], dropout=0.4, lr=$2\times10^{-4}$, weight_decay=$1\times10^{-3}$.

#### Supplementary Note 3: TRIPOD+AI checklist

Reconstructing real-world metastatic lines of therapy enables progression risk stratification in breast cancer

| Section/topic | Item | D/E | Checklist item | Reported on page number |
|---|---|---|---|---|
| **Title** | | | | |
| Title | 1 | D;E | Identify the study as developing or evaluating the performance of a multivariable prediction model, the target population, and the outcome to be predicted | Title, p. 1 |
| **Abstract** | | | | |
| Abstract | 2 | D;E | See TRIPOD+AI for Abstracts checklist | Abstract, pp. 1–2 |
| **Introduction** | | | | |
| Background | 3a | D;E | Explain the healthcare context (including whether diagnostic or prognostic) and rationale for developing or evaluating the prediction model, including references to existing models | Introduction, pp. 2–3 |
| | 3b | D;E | Describe the target population and the intended purpose of the prediction model in the context of the care pathway, including its intended users (eg, healthcare professionals, patients, public) | Abstract, pp. 1–2; Introduction, pp. 2–3; Discussion, pp. 6–11; Methods, "PFS labeling and censoring", p. 13 |
| | 3c | D;E | Describe any known health inequalities between sociodemographic groups | Not reported |
| Objectives | 4 | D;E | Specify the study objectives, including whether the study describes the development or validation of a prediction model (or both) | Introduction, pp. 2–3 |
| **Methods** | | | | |
| Data | 5a | D;E | Describe the sources of data separately for the development and evaluation datasets (eg, randomised trial, cohort, routine care or registry data), the rationale for using these data, and representativeness of the data | Results, "Cohort overview", p. 3; Results, "Learned risk ranking transfers to external cohorts", p. 5; Methods, "Cohort construction", p. 11; Methods, "External validation on the AACR Project GENIE BPC cohort", p. 17; Discussion, pp. 7, 9–10; Data Availability, p. 19 |
| | 5b | D;E | Specify the dates of the collected participant data, including start and end of participant accrual; and, if applicable, end of follow-up | Not reported |
| Participants | 6a | D;E | Specify key elements of the study setting (eg, primary care, secondary care, general population) including the number and location of centres | Introduction, pp. 2–3; Results, "Cohort overview", p. 3; Results, "Learned risk ranking transfers to external cohorts", p. 5; Data Availability, p. 19 |
| | 6b | D;E | Describe the eligibility criteria for study participants | Fig. 1b, p. 25; Methods, "Cohort construction" to "PFS labeling and censoring", pp. 11–13; Methods, "External validation on the AACR Project GENIE BPC cohort", p. 17; Supplementary Figure 3; Supplementary Tables 15, 27 and 28 |
| | 6c | D;E | Give details of any treatments received, and how they were handled during model development or evaluation, if relevant | Table 1, p. 30; Methods, "mLoT segmentation" to "Feature extraction", pp. 11–14; Discussion, pp. 7–8; Supplementary Tables 2–3, 7–8 and 11–12; Supplementary Figure 4 |

| Section/topic | Item | D/E | Checklist item | Reported on page number |
|---|---|---|---|---|
| Data preparation | 7 | D;E | Describe any data pre-processing and quality checking, including whether this was similar across relevant sociodemographic groups | Methods, "PFS labeling and censoring" and "Feature extraction", pp. 12–14; Supplementary Tables 15, 18 and 28; Supplementary Note 1 |
| Outcome | 8a | D;E | Clearly define the outcome that is being predicted and the time horizon, including how and when assessed, the rationale for choosing this outcome, and whether the method of outcome assessment is consistent across sociodemographic groups | Methods, "PFS labeling and censoring", pp. 12–13; Methods, "External validation on the AACR Project GENIE BPC cohort", p. 17; Supplementary Table 21 |
| | 8b | D;E | If outcome assessment requires subjective interpretation, describe the qualifications and demographic characteristics of the outcome assessors | Methods, "PFS labeling and censoring", p. 12; Discussion, p. 10 |
| | 8c | D;E | Report any actions to blind assessment of the outcome to be predicted | Not reported |
| Predictors | 9a | D | Describe the choice of initial predictors (eg, literature, previous models, all available predictors) and any pre-selection of predictors before model building | Methods, "Feature extraction", pp. 13–14; Supplementary Table 18; Supplementary Note 1 |
| | 9b | D;E | Clearly define all predictors, including how and when they were measured (and any actions to blind assessment of predictors for the outcome and other predictors) | Methods, "Feature extraction", pp. 13–14; Supplementary Table 18; Supplementary Note 1 |
| | 9c | D;E | If predictor measurement requires subjective interpretation, describe the qualifications and demographic characteristics of the predictor assessors | Methods, "Feature extraction", p. 13; Supplementary Note 1 |
| Sample size | 10 | D;E | Explain how the study size was arrived at (separately for development and evaluation), and justify that the study size was sufficient to answer the research question. Include details of any sample size calculation | Table 1, p. 30; Results, "Learned risk ranking transfers to external cohorts", p. 5; Supplementary Table 17 |
| Missing data | 11 | D;E | Describe how missing data were handled. Provide reasons for omitting any data | Fig. 1b, p. 25; Methods, "PFS labeling and censoring", pp. 12–13; Methods, "Feature extraction", pp. 13–14; Methods, "External validation on the AACR Project GENIE BPC cohort", p. 17 |
| Analytical methods | 12a | D | Describe how the data were used (eg, for development and evaluation of model performance) in the analysis, including whether the data were partitioned, considering any sample size requirements | Methods, "Model selection and training", p. 15; Methods, "External recalibration", pp. 16–17; Supplementary Note 2 |
| | 12b | D | Depending on the type of model, describe how predictors were handled in the analyses (functional form, rescaling, transformation, or any standardisation) | Methods, "Feature extraction", pp. 13–14; Methods, "Model selection and training", p. 15; Supplementary Note 1 |
| | 12c | D | Specify the type of model, rationale, all model building steps, including any hyperparameter tuning, and method for internal validation | Results, "Model performance and calibration", pp. 4–5; Methods, "Model selection and training", p. 15; Supplementary Table 14; Supplementary Note 2 |
| | 12d | D;E | Describe if and how any heterogeneity in estimates of model parameter values and model performance was handled and quantified across clusters (eg, hospitals, countries). See TRIPOD-Cluster for additional considerations | Results, "Learned risk ranking transfers to external cohorts", p. 5; Methods, "Evaluation" to "External validation on the AACR Project GENIE BPC cohort", pp. 15–17; Supplementary Tables 17 and 25 |

| Section/topic | Item | D/E | Checklist item | Reported on page number |
|---|---|---|---|---|
| | 12e | D;E | Specify all measures and plots used (and their rationale) to evaluate model performance (eg, discrimination, calibration, clinical utility) and, if relevant, to compare multiple models | Methods, “Evaluation”, pp. 15–16; Methods, “Risk stratification and Kaplan–Meier summaries”, pp. 17–18; Supplementary Tables 13, 23 and 25; Supplementary Figures 5–6 |
| | 12f | E | Describe any model updating (eg, recalibration) arising from the model evaluation, either overall or for particular sociodemographic groups or settings | Results, “Learned risk ranking transfers to external cohorts”, p. 5; Methods, “External recalibration”, pp. 16–17 |
| | 12g | E | For model evaluation, describe how the model predictions were calculated (eg, formula, code, object, application programming interface) | Methods, “Evaluation” and “External recalibration”, p. 16; Methods, “External validation on the AACR Project GENIE BPC cohort”, p. 17 |
| Class imbalance | 13 | D;E | If class imbalance methods were used, state why and how this was done, and any subsequent methods to recalibrate the model or the model predictions | Not applicable |
| Fairness | 14 | D;E | Describe any approaches that were used to address model fairness and their rationale | Not reported |
| Model output | 15 | D | Specify the output of the prediction model (eg, probabilities, classification). Provide details and rationale for any classification and how the thresholds were identified | Results, “Model performance and calibration”, p. 4; Results, “Prognostic stratification despite a front-loaded event distribution”, pp. 5–6; Figs. 2 and 4, pp. 26 and 28; Methods, “Risk stratification and Kaplan–Meier summaries”, pp. 17–18 |
| Training versus evaluation | 16 | D;E | Identify any differences between the development and evaluation data in healthcare setting, eligibility criteria, outcome, and predictors | Results, “Agreement of mLoT reconstruction with curated GENIE BPC regimens”, pp. 3–4; Results, “Learned risk ranking transfers to external cohorts”, p. 5; Methods, “External validation on the AACR Project GENIE BPC cohort”, p. 17; Discussion, pp. 8–10; Supplementary Tables 17, 19 and 20 |
| Ethical approval | 17 | D;E | Name the institutional research board or ethics committee that approved the study and describe the participant informed consent or the ethics committee waiver of informed consent | Not reported |
| **Open science** | | | | |
| Funding | 18a | D;E | Give the source of funding and the role of the funders for the present study | Funding, p. 20 |
| Conflicts of interest | 18b | D;E | Declare any conflicts of interest and financial disclosures for all authors | Competing Interests, p. 20 |
| Protocol | 18c | D;E | Indicate where the study protocol can be accessed or state that a protocol was not prepared | Not reported |
| Registration | 18d | D;E | Provide registration information for the study, including register name and registration number, or state that the study was not registered | Not reported |
| Data sharing | 18e | D;E | Provide details of the availability of the study data | Data Availability, p. 19 |
| Code sharing | 18f | D;E | Provide details of the availability of the analytical code | Code Availability, p. 19 |
| **Patient and public involvement** | | | | |
| Patient and public involvement | 19 | D;E | Provide details of any patient and public involvement during the design, conduct, reporting, interpretation, or dissemination of the study or state no involvement | Not reported |
| **Results** | | | | |

| Section/topic | Item | D/E | Checklist item | Reported on page number |
|---|---|---|---|---|
| Participants | 20a | D;E | Describe the flow of participants through the study, including the number of participants with and without the outcome and, if applicable, a summary of the follow-up time. A diagram may be helpful | Fig. 1b, p. 25; Table 1, p. 30; Results, "Prognostic stratification despite a front-loaded event distribution", pp. 5–6; Methods, "PFS labeling and censoring", pp. 12–13; Supplementary Tables 17 and 19a; Supplementary Figure 6 |
| | 20b | D;E | Report the characteristics overall and, where applicable, for each data source or setting, including the key dates, key predictors (including demographics), treatments received, sample size, number of outcome events, follow-up time, and amount of missing data. A table may be helpful. Report any differences across key demographic groups | Table 1, p. 30; Methods, "Feature extraction", pp. 13–14; Methods, "External validation on the AACR Project GENIE BPC cohort", p. 17; Supplementary Tables 1–3 and 17 |
| | 20c | E | For model evaluation, show a comparison with the development data of the distribution of important predictors (demographics, predictors, and outcome) | Results, "Learned risk ranking transfers to external cohorts", p. 5; Methods, "External validation on the AACR Project GENIE BPC cohort", p. 17; Supplementary Tables 17, 20 and 25 |
| Model development | 21 | D;E | Specify the number of participants and outcome events in each analysis (eg, for model development, hyperparameter tuning, model evaluation) | Tables 1–2, p. 30; Methods, "Model selection and training", p. 15; Supplementary Tables 13, 17 and 21–27 |
| Model specification | 22 | D | Provide details of the full prediction model (eg, formula, code, object, application programming interface) to allow predictions in new individuals and to enable third party evaluation and implementation, including any restrictions to access or reuse (eg, freely available, proprietary) | Code Availability, p. 19; Supplementary Tables 14 and 18; Supplementary Note 2 |
| Model performance | 23a | D;E | Report model performance estimates with confidence intervals, including for any key subgroups (eg, sociodemographic). Consider plots to aid presentation | Results, "Model performance and calibration" and "Learned risk ranking transfers to external cohorts", pp. 4–5; Fig. 2, p. 26; Table 2, p. 30; Supplementary Tables 4, 13, 17 and 23–25; Supplementary Figures 1 and 5 |
| | 23b | D;E | If examined, report results of any heterogeneity in model performance across clusters. See TRIPOD-Cluster for additional details | Results, "Learned risk ranking transfers to external cohorts", p. 5; Supplementary Tables 17 and 25b–f; Supplementary Figures 5–6 |
| Model updating | 24 | E | Report the results from any model updating, including the updated model and subsequent performance | Results, "Learned risk ranking transfers to external cohorts", p. 5; Discussion, p. 8; Supplementary Table 25d–f; Supplementary Figure 6 |
| **Discussion** | | | | |
| Interpretation | 25 | D;E | Give an overall interpretation of the main results, including issues of fairness in the context of the objectives and previous studies | Discussion, pp. 6–11 |
| Limitations | 26 | D;E | Discuss any limitations of the study (such as a non-representative sample, sample size, overfitting, missing data) and their effects on any biases, statistical uncertainty, and generalisability | Discussion, pp. 8–11 |

| Section/topic | Item | D/E | Checklist item | Reported on page number |
|---|---|---|---|---|
| Usability of the model in the context of current care | 27a | D | Describe how poor quality or unavailable input data (eg, predictor values) should be assessed and handled when implementing the prediction model | Results, "Model performance and calibration" and "Learned risk ranking transfers to external cohorts", pp. 4–5; Methods, "Feature extraction", pp. 13–14; Methods, "External validation on the AACR Project GENIE BPC cohort", p. 17; Discussion, pp. 7–8 |
| | 27b | D | Specify whether users will be required to interact in the handling of the input data or use of the model, and what level of expertise is required of users | Not reported |
| | 27c | D;E | Discuss any next steps for future research, with a specific view to applicability and generalisability of the model | Discussion, pp. 8, 10–11 |

From: Collins GS, Moons KGM, Dhiman P, et al. BMJ 2024;385:e078378. doi:10.1136/bmj-2023-078378
